# Quantifying the impact of SARS-CoV-2 temporal vaccination trends and disparities on disease control

**DOI:** 10.1101/2023.02.23.23286326

**Authors:** Sophie L. Larsen, Ikgyu Shin, Jefrin Joseph, Haylee West, Rafael Anorga, Gonzalo E. Mena, Ayesha S. Mahmud, Pamela P. Martinez

## Abstract

SARS-CoV-2 vaccines were developed and distributed during a global crisis at unprecedented speed. Still, little is known about trends in vaccine uptake over time, their association with socioeconomic inequality, and the impact of these temporal trends on disease control. By analyzing data from dozens of countries, we examined vaccination rates across high and low socioeconomic (SES) groups, showing that socioeconomic disparities in the fraction of the population vaccinated exist at both national and sub-national levels. We also identified two distinct vaccination trends: one characterized by rapid initial roll-out, quickly reaching a plateau; and another trend that is sigmoidal and slow to begin. Informed by these patterns, we implemented an SES-stratified mechanistic model, finding profound differences across the two vaccination types in the burden of infections and deaths. The timing of initial roll-out has a more significant effect on transmission and deaths than the eventual level of coverage or the degree of SES disparity. Surprisingly, the speed of the roll-out is not associated with wealth inequality or GDP per capita of countries. While socioeconomic disparity should be addressed, accelerating the initial roll-out for all groups is a broadly accessible intervention and has the potential to minimize the burden of infections and deaths across socioeconomic groups.

## Introduction

Severe acute respiratory syndrome coronavirus 2 (SARS-CoV-2) is a respiratory virus first detected in China in late 2019 and declared a pandemic in March 2020. Three years later, SARS-CoV-2 is responsible for over 650 million confirmed cases and over 6.6 million deaths worldwide [1]. Disparities have been found in coronavirus disease 2019 (COVID-19) outcomes across demographic groups (e.g. [2–4]), as well as the consequences of racial and socioeconomic disparities in infection fatality rates and all-cause mortality, [2, 5, 6], COVID-19 testing rates [2, 7, 8], transmission [9], and in the ability to social distance and reduce mobility [2, 9, 10].

This pandemic also prompted an unprecedented global development of hundreds of vaccine candidates, with around two dozen authorized for use [11]. Vaccination began in late 2020 and the number of administered vaccine doses totals over 13 billion worldwide as of February 2023, exceeding the global population [1]. Global vaccination has been estimated to have averted at least 14 million deaths [12], but there has been unequal distribution of the global vaccine supply between high-income countries and low- and middle-income countries [13, 14]. Mathematical models have been useful in understanding optimal SARS-CoV-2 vaccine prioritization strategies based on age [15] and vaccine sharing to places with lower vaccine availability [16, 17]. At the same time, recent studies have highlighted the need to incorporate racial and socioeconomic aspects of host heterogeneity into the study of infectious disease dynamics [10, 18, 19] and into vaccine prioritization [20]. While local heterogeneity in vaccine coverage has also recently been identified at limited time scales [21, 22], still little is known about the temporal trends of vaccination at the national and sub-national levels, the potential differences across socioeconomic status, and their consequences for incidence and deaths due to SARS-CoV-2 infections.

Here, we analyze weekly vaccination data from dozens countries and territories, and more than 8000 subnational locations to examine temporal dynamics of vaccination. Our results show that, at the national level, percentage of the population vaccinated is strongly associated with per capita gross domestic product (GDP), and at the sub-national level, there are persistent socioeconomic inequalities in vaccine uptake across continents. Strikingly, we also distinguish substantial variability in the speed of the initial roll-out, independent of GDP or continent. By implementing these different vaccination trends into a mechanistic model, we find that a faster vaccine roll-out has the most significant effect on the incidence and mortality than the proportion of population vaccinated in the long-term. Accelerating the initial roll-out can act as an alternative and accessible intervention for many countries with limited access to the vaccine.

## Results

### Global vaccine coverage and socioeconomic disparity

We analyzed data on vaccination and gross domestic product per capita for 160 countries and territories [23–57], finding a significant positive association between the GDP per capita of a country and the percentage of the population vaccinated against SARS-CoV-2 with at least one dose (fig. 1A). We found that as of June 2022, 97 of these countries (60%) have at least 50% of their population vaccinated, a finding that is consistent with known disparities in vaccine distribution across country income levels [13, 14]. We also collected publicly available temporal data for 34 countries and territories with a sub-national resolution, selecting those that have at least 50% population coverage by a first dose to adequately capture the temporal dynamics and with at least 12 sub-national localities reporting in the data so that individual localities do not dominate the SES-based analysis (fig. 1B).

**Figure 1:**
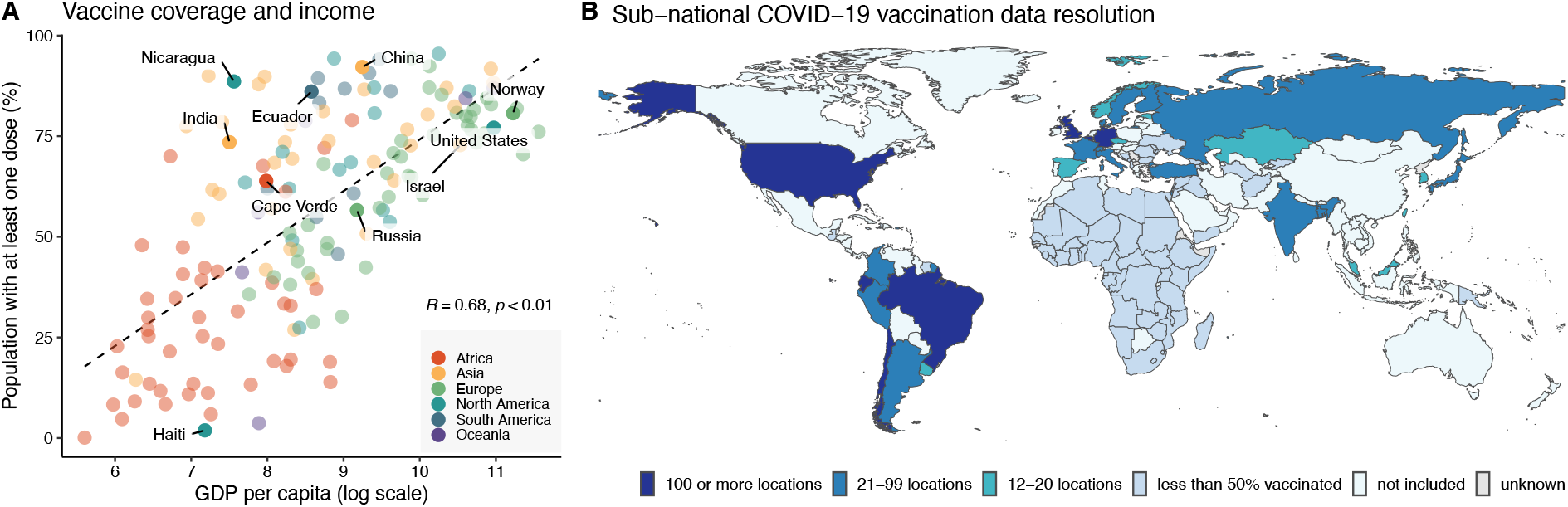
SARS-CoV-2 vaccine coverage. (A) Association between population vaccinated with at least one dose and gross domestic product (GDP) per capita for 160 countries and territories. Vaccine and population data come from [23–55] and were compiled by the Financial Times as of June 22, 2022. GDP data are sourced from World Bank [56] and PCBS [57]. (B) Sub-national vaccination data resolution. Based on publicly available data for places with 50% or more of their population vaccinated with at least one dose, we identified 7 countries/territories with vaccination rates for 100 or more locations, 17 for 21-99, and 10 for 12-20 locations. In this study, we excluded 78 countries that have greater than 50% vaccination coverage due to a lack of high-resolution sub-national data, which are referenced as ‘not included’ in the legend. Details of the data sources and dates of data collection are reported in tables S1 and S2, respectively.

From the 34 countries and territories with available temporal data, we split the localities at the sub-national level into quantiles based on measures representative of socioeconomic status (SES) such as income or poverty rates (table S1). We then analyze the vaccination temporal trends of 81 places by SES, spanning 26 countries from 5 continents, 33 states of the United States and 24 states of Brazil, and a range of national GDP per capita. We find that high SES groups are experiencing higher rates of vaccination than low SES in 83% of the places analyzed (fig. 2 and fig. S1), a disparity that persists over time in the majority of cases (fig. S2 and fig. S3). We further analyzed the degree of disparity using two metrics, the ratio and the difference for the average, the maximum, and the percentage of the population vaccinated at the final week (fig. S4-S5, table S3). Across these 6 metrics, we found that California (United States,CA), Florida (United States, FL), Colombia, Malaysia, and Israel are places that have the highest disparity, while Argentina, Amapa (Brazil, AP), Republic of Korea, and Finland have some of the least disparity observed. We also identified places with disparities that favor low over high SES groups, as is the case of Amazonas (Brazil, AM) and Nevada (United States, NV).

**Figure 2:**
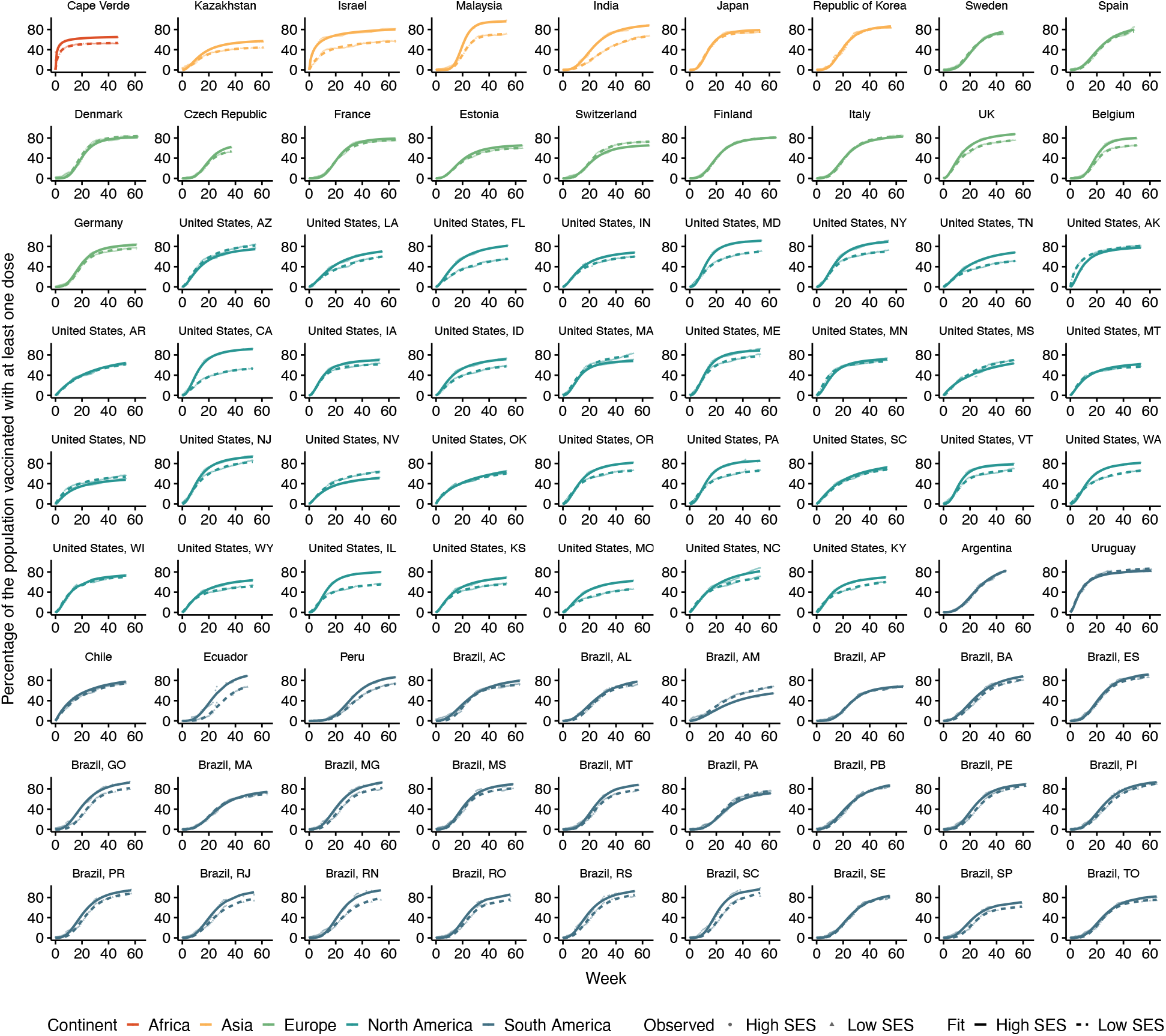
Observed (points) and predicted (lines) temporal trends of first-dose COVID-19 vaccine coverage at the sub-national level for high (solid) and low (dashed) SES groups, spanning 26 countries. A remaining 9 places are analyzed in figure S1, and full list of excluding factors are described in the supplementary text. Week 0 is the first week available in the data.

In order to evaluate possible biases of the quantile-based analysis due to the relative inequality within each location, we evaluated the degree of disparity as a function of the spatial resolution of the quantile-based analysis. We found no significant association in most pairwise comparisons across the six disparity metrics evaluated (fig. S6). We also analyzed the Lorenz curves for 83 places and estimated a corresponding Gini coefficient of vaccine inequality, finding that vaccines are unequally distributed, with a wide range of Gini values (fig. S7). Using publicly reported Gini indices of income inequality for countries/states (sources in table S6), we found a significant association between the Gini coefficients of vaccine disparity and the Gini index of income inequality (fig. S8, S9), and with the quantile-based analysis (fig. S10), suggesting that socioeconomic inequality is a likely driver of vaccine inequity across countries/states.

### Distinct vaccination temporal trends

Inspired by a method traditionally used in ecology [58], enzyme kinetics [59], and life expectancy [60], we fit a functional response *f*(*t*) to the percentage of the population vaccinated over time t (eq. 1), starting at the first week of available data for each country. This allows us to quantify for each country/state the potential maximum percentage of the population vaccinated *V*_*m*_, the week at which half of the vaccination potential is reached *W*_*h*_, and a standard parameter *k* that relates to the shape of the temporal trend.

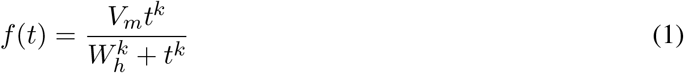

Our results not only show that the fitting successfully captures the vaccination trends in most places (lines in fig. 2), but we also identified distinct temporal dynamics that vary in shape from a very fast initial roll-out, as is the case for Cape Verde, to sigmoidal trends, like those observed in Malaysia. To further characterize these temporal trends, we fit a finite Gaussian mixture model [61] to the values of *V*_*m*_, *W*_*h*_, and *k*, identifying 2 types of dynamics shown in figure 3A: a rapid initial roll-out, and then slowing down until reaching a steady state (concave type), and a slow beginning translated into several weeks of delay in reaching half of the vaccination potential, but with the potential to reach a higher percentage of the population vaccinated (sigmoidal type).

**Figure 3:**
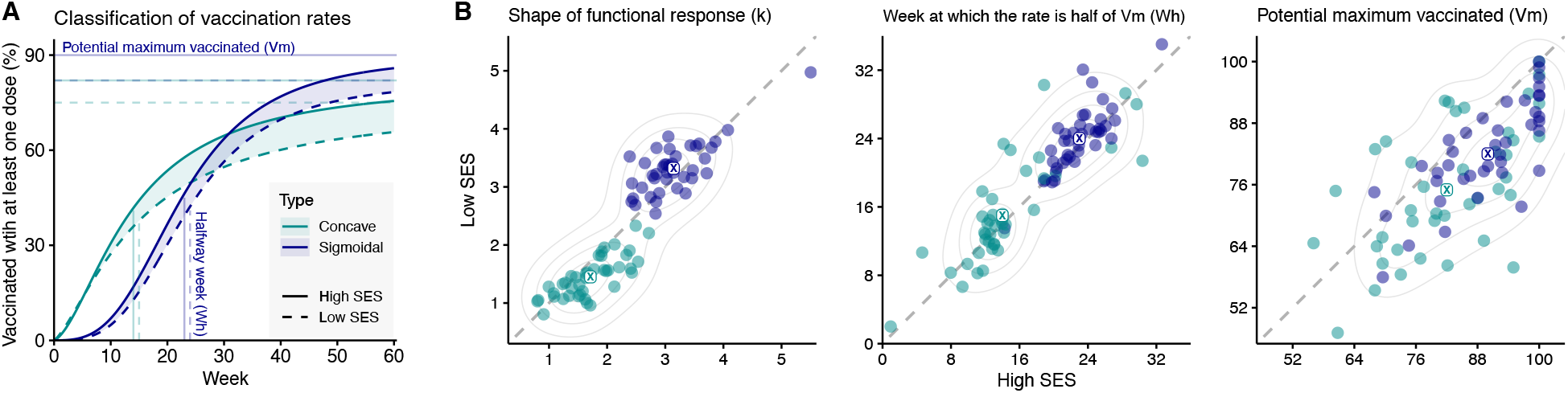
Parameters from the functional response fitting and the two temporal trends identified using the Gaussian mixture model [61]. (A) Vaccination trends over time for concave (teal) and sigmoidal (purple) types. The percentage of the cumulative population vaccinated over time for each type and SES (low in dashed line and high in solid line) were calculated using the average parameters for each type based on the classification of the 81 locations shown in figure 2. Parameters for each place can be found in table S4. (B) Values for low and high SES of the shape parameter *k*, the week at which half of the vaccination potential is reached *W*_*h*_, and of the potential maximum percentage of the population vaccinated *V*_*m*_, colored by the two temporal trends. Clustering is still apparent when displaying the United States and Brazil as countries rather than states (fig. S11).

While the cluster algorithm is informed by the three parameters, the values of *k* are the most segregated into the two types of temporal trends identified (fig. 3B). The concave type includes values of *k* with ranges of 0.8 − 2.7, while the range for the sigmoidal type has ranges of 2.4 − 5.5. While the distribution of *W*_*h*_ has some degree of overlap, the countries/states classified as sigmoidal types tend to have, on average, a halfway-week falling 8 weeks later than for the concave types, with little difference among high and low SES within each type. In contrast, the potential population vaccinated *V*_*m*_ shows a less clear partitioning, with average values of 75%, 82%, 82%, and 90%, for concave low SES, concave high SES, sigmoidal low SES, and sigmoidal high SES, respectively. When looking at the association of these two temporal classifications with GDP per capita and the Gini coefficient, *V*_*m*_ was significantly associated with wealth inequality (fig S12). Unexpectedly, we found no significant relationship between shape *k* and timing *W*_*h*_ when each variable is compared to GDP per capita and the Gini coefficient (fig. S12).

### Impact of SES and vaccination temporal trends on disease transmission

We used the outputs of the functional response fitting to calculate an average daily vaccination rate (time derivative) for the concave and sigmoidal trends over a 14 month period, finding that the concave type peaks around 3 months earlier than the sigmoidal one (fig. 4A and fig. S13). We then implemented these average daily rates in an SES-stratified SEIR model, with the vaccine intervention starting immediately or at month 7. If vaccination is started immediately, there is a larger impact, and a concave strategy is optimal for the first 12 months, until cases begin to rise (fig. 4B), likely due to the waning of immunity. Since the vaccines for SARS-CoV-2 were not available until after several months, we consider the scenario where vaccination starts at t = 7 months to be more realistic, in which a faster initial roll-out (concave type) also reduces the incidence the most (fig. 4C). For the remaining analyses, we assume the latter scenario.

**Figure 4:**
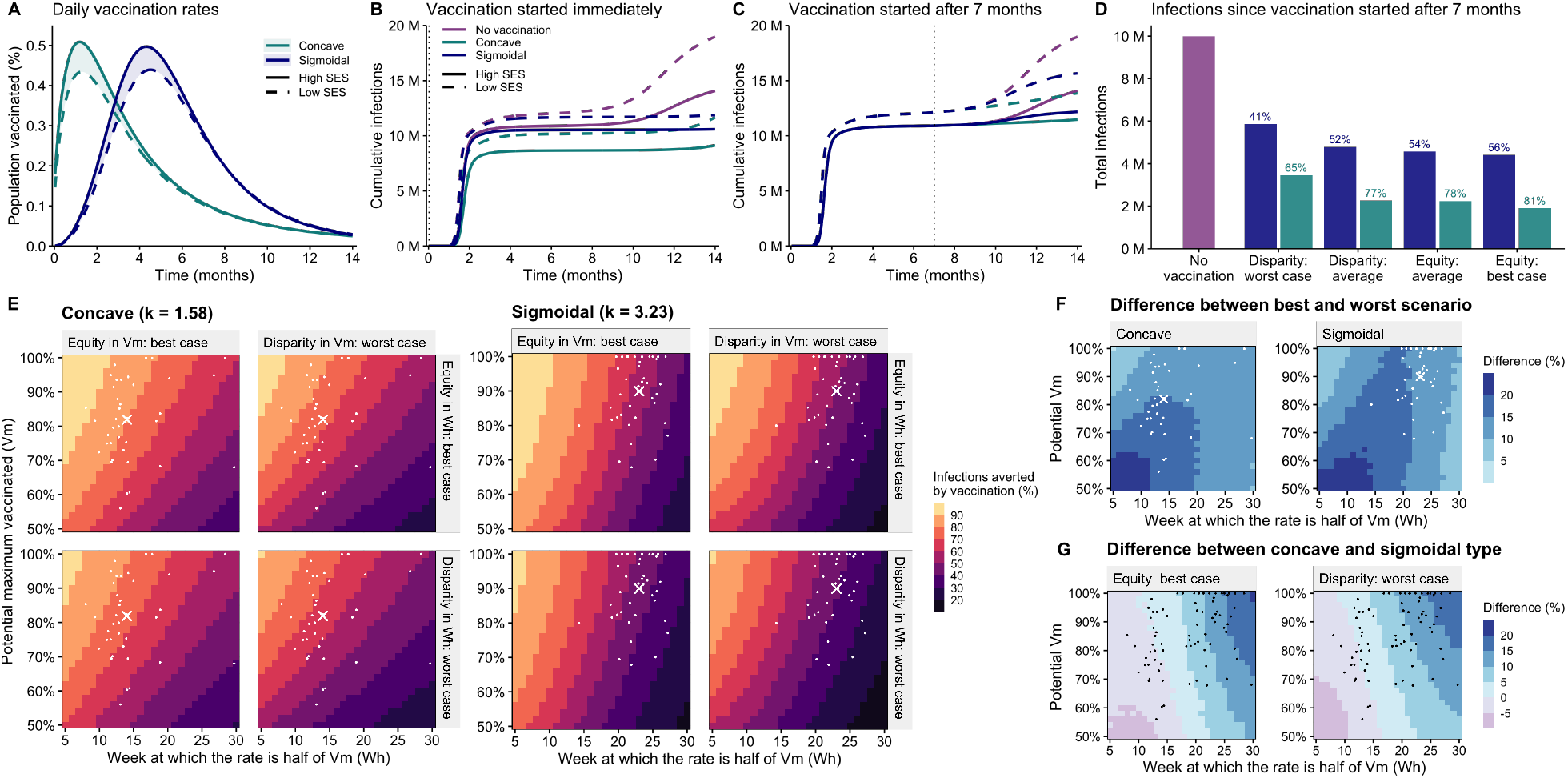
(A) Average daily vaccination rates for each type and SES group (per-country rates shown in fig. S13). In the sigmoidal type, the parameters for high SES are *Vm* = 89% and *Wh* = 23 weeks, while for low SES are *Vm* = 82.5% and *Wh* = 24 weeks. For both SES groups, *k* = 3.23, the average across SES. In the concave type, the parameters for high SES are *Vm* = 82.2% and *Wh* = 14 weeks while for low SES they are *Vm* = 75.5% and *Wh* = 15 weeks. For both SES groups, *k* = 1.58. (B) Cumulative cases per 10,000 with vaccination starting immediately. (C) Cumulative cases per 10,000 with vaccination starting at *t* = 7 months. (D) Cumulative cases per 10,000 after 14 months under 4 scenarios of sociocioeconomic disparity: disparity: average, disparity: worst case (95th percentile), equity: average (averaging high SES and low SES parameters), and equity: best case (assuming vaccination of the low SES group follow the same parameters as the high SES group. (E) Cases averted by vaccination. Keeping *k* = 1.58 for concave and *k* = 3.23 for sigmoidal, we fixed the low SES parameters at a constant disparity from high SES based on the 95th percentiles of the observed data (Δ*Vm* = 21%, Δ*Wh* = 6 weeks), or considered low SES parameters equal to high SES (equity) (fig. S16). The effect of varying parameter *k* can be found in figure S17. (F) The difference in cases averted under the best-case equity scenario and worst-case disparity, for both concave and sigmoidal types. (G) The difference in cases averted under concave versus sigmoidal types, for both best-case equity and worst-case disparity. Average high SES parameters are illustrated with a cross, while country-specific parameters for high SES are shown with dots. Model parameters are listed in table S5.

We then examined concave and sigmoidal vaccination strategies under 4 different scenarios of disparities against a no-vaccination scenario considering: (i) the average parameters for each socioeconomic group (Disparity: average), (ii) an extreme case of disparity between high and low SES (Disparity: worst case), (iii) the average parameters disregarding socioeconomic group (Equity: average), and (iv) having the low SES group be vaccinated at the same rate as the high SES group (Equity: best case). These results show that a substantial percentage of cases can be averted by moving from the worst-case disparity to any other scenario, when compared to a no-vaccination case (fig. 4D), with a reduction of up to 16% in the ‘Equity: best case’ strategy. Moreover, we find that a faster initial roll-out is always better to avert cases, reflected in an additional 24-25% of cases averted when moving from sigmoidal to concave type. This effect is slightly stronger for the mortality case, with 25-27% of deaths averted (fig. S14D).

Our findings suggest that both the temporal trend and the disparity can have a substantial impact on the outcomes. However, the effects of changes in individual parameters cannot be isolated. To this end, we independently varied *W*_*h*_ for high SES from 5 to 30 weeks and the *V*_*m*_ from 50% to 100%, fixing a constant disparity in the parameters for the ‘disparity: worst-case’ and ‘equity: best case’ scenarios (fig. 4E). Intuitively, no matter the type of concave or sigmoidal dynamics, having a higher potential of people vaccinated with an earlier halfway week is always better for both cases (fig. 4E) and deaths (fig. S14E). We also found the halfway week has a stronger effect on the outcomes than *V*_*m*_. For instance, if we analyze the equity scenario for both *W*_*h*_ and *V*_*m*_ (top-left corner in figure 4E), the cases averted could be as low as 60% (concave) or 40% (sigmoidal) if the halfway week is 30, even if the maximum vaccinated is 100%. In contrast, if we look at *V*_*m*_ = 50% and *W*_*h*_ = 5, the cases averted are around 80% in both concave and sigmoidal types.

When comparing across types, the sigmoidal dynamics are more sensitive to the values of *W*_*h*_, where the outcomes are worst at higher values of *W*_*h*_, compared to the concave, illustrated in the 20% difference in cases averted when *V*_*m*_ = 100% and *W*_*h*_ = 30. The association of a lower halfway week in countries/states classified as concave may contribute to why concave dynamics are optimal in figure 4D. When we estimated the infections averted for the values of *W*_*h*_ and *V*_*m*_ obtained from the data fitting (illustrated by the white dots), we found that the majority of places with concave vaccination temporal trends would lead to 70%-90% of infections averted, compared to a majority ranging from 50% to 70%, in the case of sigmoidal trends.

To further understand the impact of socioeconomic disparity, we compared the outcomes of the best-case equity and worst-case disparity scenarios, at a given set of *V*_*m*_ and *W*_*h*_ values (fig. 4F). We find that difference is minimal under a fast initial roll-out (*W*_*h*_ = 5) and a max. coverage (*V*_*m*_ = 100%), reflected in only a 5% and 6% difference between best-case equity and worst-case disparity, for concave and sigmoidal, respectively. Counter-intuitively, a low coverage (*V*_*m*_ = 50% − 64%) together with a fast roll-out (*W*_*h*_ =5 − 14) could generate the biggest gap between equity and disparity scenarios. However, this scenario seems to be unlikely as it isn’t represented in the parameter combinations estimated from the data (white dots). These analyses also show that while the values obtained from fitting the data fall in areas of the parameter space with low differences between both scenarios, socioeconomic disparity could lead to higher differences in the cases averted under the sigmoidal type, reflected in more areas with a difference greater than 15%.

Finally, when comparing across the two types, the results show that the concave dynamics are optimal for most of the parameter space, independent of the socioeconomic disparity (fig. 4G), and with the biggest difference observed at *W*_*h*_ = 30 and *V*_*m*_ = 100%. However, if the initial roll-out is unusually fast (e.g. *W*_*h*_ = 5 weeks), sigmoidal becomes optimal. Under these extreme conditions, sigmoidal and concave trends are very similar up to the halfway week, when sigmoidal rapidly overtakes concave and reaches the potential maximum vaccinated relatively fast (fig. S15). This further supports that a fast roll-out in the early weeks of vaccination drives a higher percentage of cases averted. Still, the area of the parameter space where sigmoidal is optimal results in only a 5-10% difference in the cases averted, indicating that the effect is weak (Fig. 4G). Similar results are shown for deaths averted (Fig. S14).

## Discussion

By analyzing publicly available data at the national and sub-national level, we examine different aspects of temporal trends in vaccination with a first dose against SARS-CoV-2, the association with socioeconomic disparities, and the impact on overall transmission and mortality. Socioeconomic and racial disparities have been previously identified in SARS-CoV-2 incidence [2, 18, 62] and in vaccination in a few places [21, 22]. We validated these findings at the global level by showing a significant association between the fraction of the population vaccinated with at least one dose and gross domestic product per capita, in line with prior observation that the global distribution of vaccines has been highly heterogeneous [13, 17].

While the impact of SARS-CoV-2 vaccination on disease outcomes has been previously studied (e.g. [12, 15, 16]), less understood has been the extent of heterogeneity in vaccination temporal trends and its consequences for disease outcomes. To this end, we characterized weekly vaccination uptake of 81 locations, disaggregated by SES, finding that most countries and states experience inequity in vaccine administration in favor of high SES groups, which do not resolve over time. Additionally, the temporal trends vary widely in shape and speed of the vaccination roll-out, which we classified as concave or sigmoidal types. We showed that concave dynamics tend to have a faster initial roll-out, which is reflected in a higher number of cases and deaths averted due to vaccination. Critically, the timing of vaccination is not associated with wealth inequality or GDP per capita, where, for example, Cape Verde has implemented a faster timing compared to many high GDP countries such as Belgium, which experienced a very slow initial roll-out. These findings suggest a broadly accessible intervention where the timing rather than allocation of vaccines can be modified early on, providing a broad benefit to the population, even when the entire population cannot be vaccinated due to limited stock. We also show that speeding up the timing of vaccine roll-out can be as effective as modifying the socioeconomic disparity, which may be more challenging to address in the short term due to structural inequities in factors such as healthcare or transportation, especially given our finding that wealth inequality is correlated with vaccine disparity.

Our paper should be considered in light of the following limitations. Previous work has shown that for influenza, association between SES and vaccination is often dependent on the SES measure selected [63]. This study is limited by the availability of vaccine and socioeconomic data at a high spatial resolution for countries, especially on the African continent. It is therefore not possible to know with certainty what happens at the individual level, leading to the potential for ecological fallacy. We analyzed first dose coverage but trends may differ for second dose or booster. First dose coverage may vary in quality as a long-term vaccination strategy depending on the quality of immune response [64]. While we did not study the causes of socioeconomic heterogeneity in vaccination, access and intent to vaccinate can vary widely across different countries, and for the case of SARS-CoV-2, it has been linked to demographic factors including income, race, political affiliation, and education [21, 65–68]. This analysis is also agnostic to age structure in the population and this limits the quantification of overall burden. Finally, we used a mechanistic SEIR model that did not consider non-pharmaceutical interventions or pathogen evolution, which could impact intent to vaccinate.

To summarize, we have analyzed the temporal trends of vaccination from thousands of places across 5 continents showing not only that vaccine inequity exists within and between countries, but also that the speed of the roll-out can play a crucial role in the impact on disease incidence, and that surprisingly, this speed is not associated with socioeconomic metrics. By promoting faster initial vaccine uptake, our work outlines concrete steps that can be taken to reduce the impacts of inequitable vaccination on the burden of disease for everyone.

## Methods

### Data collection and processing

We collected national statistics on the percentage of the population with at least one dose from [23–55], which were compiled by the Financial Times, and GDP per capita for these nations from the World Bank [56] and Palestinian Central Bureau of Statistics (PCBS) [57]. We also collected publicly available on vaccination, SES, and population size at the sub-national level from countries, territories, and states that have at least 12 locations with both vaccination and SES data to avoid single locations having an outsize impact on the dynamics. We excluded countries where less than 50% of the total population had received a first dose, to adequately capture the temporal variation across trends. Sources and dates of collection are listed in tables S1 and S2 respectively. Within each country and state, locations were grouped into quantiles by relative SES. For countries with 12-20 locations, we used 3 groups; for 21-99 we used 5 groups; for 100 or more we used 10 groups. We then computed the average proportion vaccinated at each week, for the highest and lowest SES groups in each country.

Some of the vaccination data have inconsistent reporting and apparent adjustments to the data over time. Issues with data reporting have been flagged by other authors studying vaccination data in the US [22]. We discarded data points where the cumulative vaccination was not monotonically increasing by applying the despike() function in R to the final aggregated data, which tracks a window of the median in the data and removes any points outside a specified deviation from that median.

The daily vaccination rates, estimated as an approximation to the first derivative of the fit data, is given by the following expression:

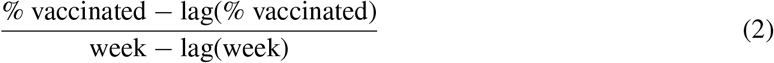

### Mechanistic model

We implemented a mechanistic SEIR model of transmission, varying the force of infection, proportion with mild or asymptomatic disease, fatality rate, and vaccination rate by socioeconomic status. Parameter values can be found in table S5. We used the functional response to estimate daily vaccination rates *ν*(*t*) for both high and low SES under two scenarios of parameter *k*, reflecting the average values of the two clusters identified in the results shown in figure 3. The following equations were implemented for the low SES group, and an identical set were implemented for high SES, but changing the parameter values:

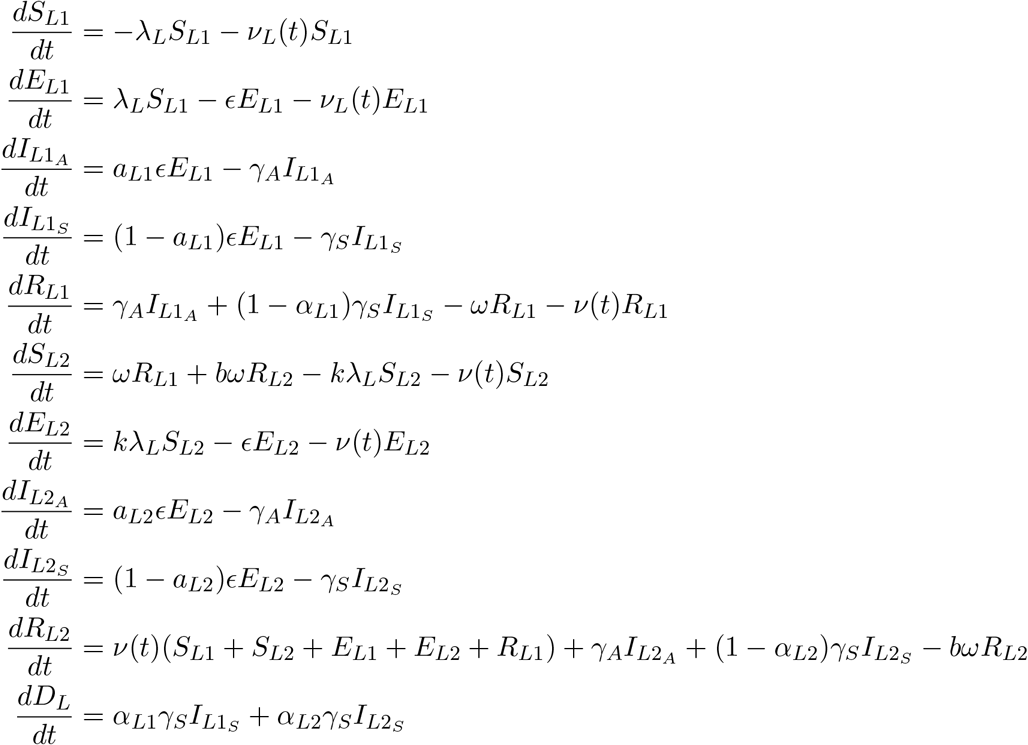

In this SEIR model, individuals start in the *S*_1_ class, and become exposed (*E*_1_) through contact with infectious individuals. Once they become infectious, they can either develop a mild 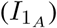 or severe 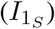 disease. The force of infection weights mild and severe infections equally, based on the observation that mild infections are substantial drivers of transmission [69]. Infectious individuals can move to the recovery class *R*_1_ with rate γ, and only individuals with severe infections can die with probability *α*. We estimated infection fatality rate (IFR) for low versus high SES as a weighted average, using the age-based estimates from Santiago, Chile [2] and United Nations population structure data for Chile [70]. We further adjusted to account for the proportion of mild/asymptomatic cases in the model, since only severe cases can die. We ran simulations with a population of 10 million low SES and 10 million high SES individuals, and assuming one exposed individual in each SES group at time 0.

Immunity from the primary infection wanes after 4 months, when individuals move from *R*_1_ to *S*_2_. Individuals in non-infected categories can also move directly to *R*_2_ through a vaccine intervention. Individuals in *R*_2_ wane their immunity after 8 months. We assume that individuals in the second compartments are less susceptible and less infectious, and there is a slightly higher proportion of mild infections, using the CDC upper bound on asymptomatic cases for both SES groups [71]. We scaled the infection fatality rate for the 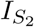 class using estimates of vaccine efficacy and waned vaccine immunity against severe disease [72, 73]. Literature on contact patterns across socioeconomic groups is limited. However, it is known that workplace and housing environments differ for low and high SES [74, 75], SES groups tend to have more mobility within-group than outside of their group [9, 76], and low SES groups have not been as able to reduce their mobility during the pandemic (e.g. [2, 10]). For these reasons, we assumed contact rates were asymmetric across SES groups. All parameter values are listed in table S5.

The force of infection λ, defined as the infection risk per susceptible individual, is given by:

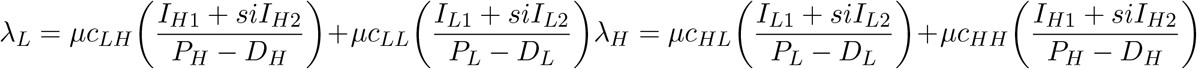

Where *μ* is the probability of infection given contact, *c*_*ij*_ is the contact rate among SES groups, and *si* is a scale parameter that reduces the infectiousness of individuals that are in the second Infected class *I*_2_.

## Data Availability

All data collected in this project are publicly available, with sources listed in the supplementary materials. All code and data to reproduce the figures in the main text will be provided upon acceptance.

## Acknowledgements

We thank the many agencies with publicly available vaccine and socioeconomic data for making it possible to conduct this analysis.

## Author Contributions

S.L.L, A.S.M., and P.P.M. conceptualized the study. S.L.L. and H.W. conducted the literature review. S.L.L, I.S., J.J., and R.A. collected and formatted the data. S.L.L. and P.P.M. analyzed the data, designed and coded the model, and wrote the manuscript. A.S.M. assisted with model design. G.E.M. assisted with the analysis. All authors edited the manuscript.

## Competing Interests

The authors declare no competing interests.

## Notes

### Competing Interest Statement

The authors have declared no competing interest.

### Funding Statement

This study did not receive any funding.

### Author Declarations

All data was available to the public before the initiation of the study. Sources can be found in the supplementary materials.

