## Supplementary Materials for "Quantifying the impact of SARS-CoV-2 temporal vaccination trends and disparities on disease control"

### S1 Supplementary text

#### S1.1 Data excluded

After considering states from Brazil and the US as separate locations, we initially analyzed 90 locations. We excluded 9 places (shown with an asterisk next to their names in figure S1) due to the following reasons: (i) data for Colombia and Puerto Rico do not differentiate between the number of doses, and thus, we cannot distinguish a person getting a second dose or someone getting their first dose; (ii) Bangladesh was still experiencing the exponential phase of the vaccination rate, so we were not able not to fit the functional response; (iii) Palestine, Turkey, Russia, and Taiwan were excluded due to insufficient weeks of data to fit; (iv) United States, AL had a large jump in the data at week 40, so we were not able to fit the functional response, and (v) Norway was excluded because the 12 locations in the data include Svalbard, which has a small population and limited socioeconomic data.

#### S1.2 Lorenz Curve and Gini Coefficient

To analyze inequity at the global scale, we compared the Gini coefficient for 58 countries and US states to the parameters from the functional response as well as disparities in the predicted data (fig. S10). Additional data sources for this analysis are listed in table S6.

To further explore disparities within countries, for places where we had population data and raw counts of vaccination, we used the disaggregated data to fit Lorenz curves. We selected the final week in the data and compared the cumulative number of vaccines administered with the corresponding population density in R, using Lc() from the Ineq package and Gini() from the DescTools package.

#### S1.3 Compartmental diagram

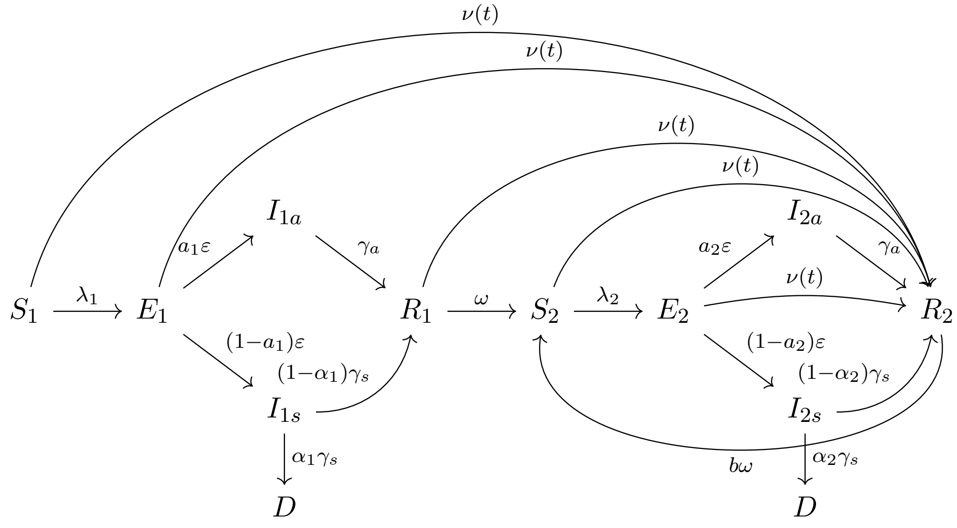


### S2 Supplementary Tables

| Country Data Sources | | | |
|---|---|---|---|
| Country | Category | Type | Link |
| Cape Verde | Vaccine | No. of adults vaccinated with 1 doses | Govt. of Cape Verde |
| | Population | municipal population (2015) | National Institute of Statistics, Cape Verde |
| | SES | poverty incidence by municipality (2007) | National Institute of Statistics, Cape Verde |
| Kazakhstan | Vaccine | number of people vaccinated with 1 component | Qaz Content |
| | Population | population (2014) | Azhgaliyeva et al. (2017) |
| | SES | poor (poverty %) (2014) | Azhgaliyeva et al. (2017) |
| Israel | Vaccine | accumulated vaccination first dose | Data Gov |
| | SES | average monthly salary of salaried employees (NIS) All employees (2019) | Central Bureau of Statistics |
| | Population | population in localities, by population group, end of 2019 | Central Bureau of Statistics |
| Palestine | Vaccine | the proportion of the vaccinated | https://corona.ps/ |
| | SES | Average Daily Wage in NIS for Wage Employees Aged 15 Years and Above in Palestine by Region and Governorate and Sex, 2020 | PCBS |
| Turkey | Vaccine | people vaccinated | Sociepy Github |
| | Population | population | Sociepy Github |
| | SES | per capita GDP, 2019 (by provinces) | TurkStat |
| Malaysia | Vaccine | cumul partial | CITF-Malaysia |
| | Population | population ('000), year 2020 | data.gov.my |
| | SES | incidence of hardcore poverty (2012) | data.gov.my |
| India | Vaccine | dose1 | Co-WIN |
| | Population | population | Sociepy GitHub |
| | SES | 2011-12 poverty rate | Reserve Bank of India |
| Japan | Vaccine | Vaccination status as 1st dose completed | Digital Agency |
| | Population | total population (2019) | e-Stat |
| | SES | 2 Yearly Income (2019) | e-Stat |
| Korea | Vaccine | firstCnt | KDX |
| | SES | gross regional income per capita (2021) | KOSIS |
| Bangladesh | Vaccine | 1st doses | MIS, DGHS |

| | | | |
|---|---|---|---|
| | Population<br>SES | population (extracted August 2022)<br>2011 estimates of mean DHS wealth index score | MIS, DGHS<br>WorldPop |
| Sweden | Vaccine<br>Population<br>SES | people vaccinated<br>population<br>at risk of poverty rate (2020) | Sociepy GitHub<br>Sociepy GitHub<br>Statistics Sweden |
| Spain | Vaccine<br>Population<br>SES | people vaccinated<br>population<br>disposable income per capita (2014) | Sociepy GitHub<br>Sociepy GitHub<br>OECD |
| Russia | Vaccine<br>Population<br>SES | people vaccinated<br>population<br>The average monthly nominal accrued wages of employees for the full range of organizations in the economy as a whole for the subjects of the Russian Federation since 2018, rubles (2021) | Sociepy GitHub<br>Sociepy Github<br>Rosstat |
| Denmark | Vaccine<br>SES | total coverage 1 dose<br>primary income (2020) | Statens Serum Institut<br>StatBank |
| Czech Republic | Vaccine<br>Population<br>SES | people vaccinated<br>population<br>GDP per capita (2020) | Sociepy GitHub<br>Sociepy GitHub<br>CZSO |
| France | Vaccine<br>Population<br>SES | dose1<br>population (2019)<br>median standard of living (2018) | data.gouv.fr<br>Insee<br>Insee |
| Estonia | Vaccine<br>SES | population coverage (MeasurementType = vaccinated, VaccinationSeries = 1)<br>equalized yearly disposable income by county and sex (2020) | TEHIK<br>andmed.stat.ee |
| Switzerland | Vaccine<br>SES | COVID19 At Least One Dose Persons<br>social assistance rate (Ind 11 01) (2019) | opendata.swiss<br>Federal Statistical Office |
| Finland | Vaccine<br>Population<br>SES | First dose<br>population (2021)<br>Social assistance long-term recipient persons during year as % of total population (id:4021), 2020 | Finnish institute for health and welfare<br>Finnish institute for health and welfare<br>Finnish institute for health and welfare |
| Norway | Vaccine | dose 1 | FHI |

| | | | |
|---|---|---|---|
| Norway (except Svalbard) | SES | Total income, median (NOK) - 2020 | SSB |
| | Population | Population of Norway 2020, by county | SSB |
| Norway (Svalbard) | SES | average gross income (NOK), 2006 (adjusted for inflation to 2020) | SSB |
| | SES | inflation adjustment to 2020 NOK | CPI Inflation Calculator |
| | Population | population 2021 | SSB |
| Italy | Vaccine | d1 + dpi (single dose and single dose after illness) | Developers Italia GitHub |
| | Population | population by regions (2020) | Istat |
| | SES | net wealth per capita (2019) | Banca d'Italia |
| UK | Vaccine | cum Vaccination First Dose Up take By Vaccination Date Percentage | gov.UK |
| | SES | income deprivation rate (2019) | Office for National Statistics |
| Belgium | Vaccine | cumul (dose = A) | Sciensano |
| | Population | population (2021) | StatBel |
| | SES | total net taxable income (converted to per capita) (2019) | StatBel |
| Germany | Vaccine | Impfschutz = 1 | Robert Koch Institute GitHub |
| | SES | GISD 2012 | GESIS |
| | Population | population | opendatasoft |
| Puerto Rico | Vaccine | nu dosis | Departamento de Salud |
| | Population | Population estimates, July 1, 2019, (V2019) | US Census Bureau |
| | SES | Median household income (in 2019 dollars), 2015-2019 | US Census Bureau |
| United States | Vaccine | Percent of 18+ Pop with at least one Dose by State of Residence | CDC |
| | SES | median household income (2019) | USDA |
| Argentina | Vaccine | people vaccinated | Sociepy GitHub |
| | Population | population | Sociepy GitHub |
| | SES | Average remuneration of registered workers in the private sector (2017) | Ministerio de Trabajo, Empleo, o Seguridad Social |
| Uruguay | Vaccine | people vaccinated | 3dgiordano GitHub |
| | SES | Median household income (no rental value and no bonus, at current prices), according to departments (2020) | INE |
| | Population | Population counted (corrected for absent residents). 2011 Census | INE |
| Colombia | SES | poverty index (2020) | DANE |

| | | | |
|---|---|---|---|
| | Vaccine<br>Population | applied dose<br>population (2020) | gov.co<br>DANE |
| Peru | Vaccine<br><br>Location<br><br>Population<br>SES<br>Locations<br>Locations | dosis = 1<br><br>vaccination centers<br><br>population (2020)<br>total poverty %<br>ubigeo codes<br>ubigeo codes | Plataforma Nacional de Datos Abiertos<br>Plataforma Nacional de Datos Abiertos<br>RENIEC<br>ubigeo GitHub package<br>ubigeo GitHub package<br>Plataforma Nacional de Datos Abiertos |
| Ecuador | Vaccine<br>SES | primera dosis<br>Incidence of poverty estimation (2014) | Andrab S.A. GitHub<br>INEC/World Bank |
| Brazil | Vaccine<br>SES<br><br>Population<br><br>Location | 1a dose, dose, unica<br>population in extreme poverty (2009)<br>population (2009)<br><br>municipal codes | openDataSUS<br>Brazilian Institute of Geography and Statistics<br>Brazilian Institute of Geography and Statistics<br>datasets-br GitHub |
| Chile | SES<br><br>Vaccine | mean income (2017)<br><br>primera dosis | Ministry of Social Development and Family<br>MinCiencia GitHub |
| Wayback Machine | Reference | used archived pages of Kazakhstan and Palestine to construct time series | Internet Archive |

Table S1: Sources for each country's vaccine and socioeconomic data.

| Country Timepoints | |
|---|---|
| Country/Territory | Most Recent Date |
| Turkey | 8/25/2021 |
| Russia | 9/11/2021 |
| Czech Republic | 9/19/2021 |
| Sweden | 11/7/2021 |
| Belgium | 12/13/2021 to 12/19/2021 |
| Spain | 12/16/2021 |
| Argentina | 12/16/2021 |
| Puerto Rico | 12/26/2021 |
| Norway | 12/27/2021 |
| Ecuador | 12/31/2021 |
| UK | 1/2/2022 |
| United States | 1/12/2022 |
| Bangladesh | 2/27/2022 |
| Malaysia | 3/5/2022 |
| Chile | 3/6/2022 |
| France | 3/30/2022 |
| Germany | 4/3/2022 |
| Taiwan | 4/11/2022 |
| Republic of Korea | 4/11/2022 |
| Japan | 4/21/2022 |
| Estonia | 5/2/2022 |
| Switzerland | 5/2/2022 |
| Finland | 5/3/2021 to 5/9/2021 |
| Uruguay | 5/4/2022 |
| Colombia | 5/4/2022 |
| Brazil | 5/24/2022 to 5/25/2022 |
| Italy | 5/25/2022 |
| Palestine | 5/31/2022 |
| Israel | 6/12/2022 |
| Cape Verde | 6/12/2022 |
| Kazakhstan | 6/17/2022 |
| India | 6/18/2022 to 6/24/2022 |
| Denmark | 6/21/2022 |

Table S2: Summary of the most recent timepoints in each country's vaccination dataset.

| Disparity Rankings | | | | | | | | |
|---|---|---|---|---|---|---|---|---|
| Name | Ratio | | | Delta | | | Gini(wealth) | Gini(vax) |
| | Final | Max | Avg | Final | Max | Avg | | |
| Colombia | 1 | 33 | 65 | 1 | 1 | 20 | 51.3 | NA |
| United States, CA | 2 | 23 | 3 | 2 | 2 | 1 | 49 | 51.12 |
| Turkey | 3 | 17 | 2 | 8 | 8 | 9 | 41.9 | NA |
| United States, FL | 4 | 8 | 6 | 3 | 7 | 3 | 49 | 47.9 |
| Israel | 5 | 7 | 4 | 5 | 3 | 2 | 39 | 51.54 |
| United States, IL | 6 | 24 | 13 | 7 | 9 | 8 | 48 | 53.1 |
| Malaysia | 7 | 34 | 11 | 4 | 6 | 5 | 41.1 | 29.60 |
| United States, MO | 8 | 50 | 14 | 14 | 17 | 12 | 46.32 | 58.12 |
| United States, TN | 9 | 14 | 21 | 15 | 18 | 17 | 47.86 | 55.02 |
| Ecuador | 10 | 1 | 1 | 9 | 4 | 7 | 45.7 | 61.69 |
| Brazil, RN | 11 | 19 | 8 | 6 | 11 | 21 | 53 | 59.25 |
| India | 12 | 26 | 5 | 10 | 5 | 4 | 35.7 | 63.15 |
| United States, MD | 13 | 36 | 17 | 11 | 10 | 6 | 45.13 | 36.18 |
| Norway | 14 | 69 | 34 | 17 | 20 | 30 | 27.6 | NA |
| Kazakhstan | 15 | 39 | 20 | 30 | 32 | 29 | 27.8 | 25.32 |
| United States, NY | 16 | 25 | 30 | 12 | 14 | 14 | 51.02 | 41.13 |
| United States, PA | 17 | 56 | 29 | 13 | 12 | 10 | 46.8 | 49.67 |
| United States, ID | 18 | 12 | 16 | 22 | 26 | 13 | 44.57 | 52.98 |
| United States, WA | 19 | 61 | 38 | 20 | 16 | 18 | 45.6 | 50.13 |
| United States, OR | 20 | 16 | 37 | 19 | 23 | 22 | 46 | 43.69 |
| United States, NC | 21 | 31 | 52 | 16 | 19 | 39 | 47.48 | 57.18 |
| Cape Verde | 22 | 35 | 22 | 28 | 22 | 11 | 42.4 | 48 |
| United States, KS | 23 | 13 | 35 | 26 | 33 | 34 | 45.55 | 56.62 |
| Belgium | 24 | 46 | 25 | 23 | 24 | 26 | 27.2 | 46.30 |
| Brazil, SC | 25 | 27 | 9 | 18 | 15 | 15 | 42 | 57.42 |
| United States, WY | 26 | 6 | 27 | 33 | 38 | 40 | 43 | 38.91 |
| Brazil, GO | 27 | 18 | 7 | 21 | 13 | 16 | 49 | 60.72 |
| Russia | 28 | 21 | 10 | 61 | 59 | 57 | 37.5 | NA |
| Peru | 29 | 45 | 26 | 24 | 25 | 25 | 41.5 | 52.40 |
| United States, LA | 30 | 20 | 33 | 35 | 35 | 33 | 49.03 | 47 |
| Czech Republic | 31 | 32 | 41 | 42 | 48 | 53 | 25 | 27.07 |
| Brazil, RJ | 32 | 29 | 15 | 29 | 21 | 35 | 52 | 54.57 |
| United States, KY | 33 | 11 | 31 | 37 | 30 | 36 | 47.41 | 66.29 |
| UK | 34 | 66 | 44 | 27 | 36 | 24 | 35.1 | NA |
| Brazil, MG | 35 | 40 | 18 | 25 | 29 | 28 | 50 | 72.31 |
| Palestine | 36 | 75 | 47 | 39 | 43 | 23 | 33.7 | NA |
| Brazil, RO | 37 | 57 | 43 | 31 | 37 | 43 | 46 | 55.54 |
| United States, VT | 38 | 51 | 39 | 34 | 28 | 19 | 44 | 43.4 |
| United States, IN | 39 | 71 | 55 | 43 | 45 | 45 | 44.94 | 55.74 |
| Brazil, TO | 40 | 70 | 60 | 36 | 46 | 56 | 50 | 61.68 |

| | | | | | | | | |
|---|---|---|---|---|---|---|---|---|
| United States, ME | 41 | 74 | 53 | 32 | 34 | 31 | 45 | 39.24 |
| United States, IA | 42 | 4 | 36 | 44 | 49 | 42 | 44 | 59.13 |
| Brazil, SP | 43 | 28 | 12 | 49 | 42 | 38 | 53 | 68.72 |
| Brazil, RS | 44 | 47 | 24 | 38 | 39 | 32 | 49 | 65.57 |
| Brazil, MS | 45 | 73 | 56 | 41 | 47 | 50 | 48 | 55.28 |
| Brazil, AC | 46 | 10 | 19 | 47 | 51 | 49 | 56 | 45.4 |
| United States, NJ | 47 | 49 | 50 | 40 | 31 | 27 | 47.82 | 24.15 |
| Germany | 48 | 48 | 48 | 48 | 53 | 46 | 31.9 | 39.14 |
| Brazil, MT | 49 | 53 | 42 | 50 | 41 | 41 | 47 | 60.61 |
| Puerto Rico | 50 | 58 | 54 | 45 | 50 | 61 | NA | NA |
| Brazil, PR | 51 | 44 | 23 | 46 | 27 | 37 | 49 | 66.59 |
| Brazil, BA | 52 | 62 | 46 | 51 | 52 | 47 | 60 | 70.19 |
| Brazil, AL | 53 | 52 | 49 | 54 | 54 | 55 | 53 | 60.58 |
| Estonia | 54 | 72 | 57 | 58 | 58 | 52 | 30.3 | NA |
| United States, OK | 55 | 55 | 70 | 59 | 62 | 67 | 46.52 | 56.3 |
| United States, MT | 56 | 64 | 72 | 60 | 64 | 69 | 45.87 | 39.61 |
| Spain | 57 | 67 | 62 | 53 | 56 | 59 | 34.7 | 29.05 |
| Bangladesh | 58 | 54 | 68 | 55 | 60 | 78 | 32.4 | NA |
| Brazil, PI | 59 | 60 | 40 | 52 | 44 | 48 | 54 | 59.24 |
| United States, SC | 60 | 3 | 32 | 56 | 61 | 54 | 46.9 | 37.81 |
| United States, MN | 61 | 81 | 80 | 62 | 67 | 73 | 44.9 | 57.41 |
| Brazil, PE | 62 | 42 | 28 | 57 | 40 | 44 | 56 | 64.01 |
| Brazil, ES | 63 | 76 | 69 | 63 | 65 | 58 | 51 | 44.98 |
| Japan | 64 | 82 | 76 | 64 | 71 | 64 | 32.9 | 40.69 |
| Chile | 65 | 77 | 58 | 66 | 55 | 51 | 44.4 | 41.24 |
| United States, AR | 66 | 2 | 45 | 69 | 72 | 63 | 47 | 53.73 |
| Brazil, SE | 67 | 68 | 59 | 65 | 73 | 68 | 56 | 56.6 |
| France | 68 | 80 | 74 | 67 | 63 | 60 | 32.4 | 32.26 |
| Brazil, MA | 69 | 63 | 73 | 68 | 74 | 71 | 54 | 70.36 |
| United States, WI | 70 | 30 | 67 | 70 | 70 | 66 | 44 | 54.78 |
| Sweden | 71 | 83 | 83 | 71 | 78 | 77 | 30 | 40.16 |
| Brazil, PB | 72 | 43 | 51 | 72 | 68 | 62 | 56 | 62.98 |
| Brazil, AP | 73 | 37 | 75 | 73 | 79 | 75 | 59 | 30.61 |
| Taiwan | 74 | 84 | 71 | 74 | 77 | 65 | 33.6 | NA |
| Republic of Korea | 75 | 86 | 77 | 75 | 84 | 74 | 31.4 | 40.46 |
| Finland | 76 | 41 | 66 | 76 | 75 | 72 | 27.3 | NA |
| Argentina | 77 | 59 | 63 | 77 | 69 | 70 | 42.9 | 49.14 |
| Denmark | 78 | 85 | 82 | 78 | 87 | 80 | 28.2 | NA |
| Italy | 79 | 38 | 61 | 79 | 66 | 76 | 35.9 | 29.73 |
| Brazil, PA | 80 | 79 | 81 | 80 | 80 | 81 | 53 | 65.34 |
| United States, AK | 81 | 88 | 88 | 81 | 90 | 87 | 41.74 | 45.43 |
| Uruguay | 82 | 78 | 78 | 82 | 85 | 79 | 39.7 | 45.19 |
| United States, AZ | 83 | 90 | 85 | 85 | 86 | 84 | 46.82 | 31.75 |

| | | | | | | | | |
|---|---|---|---|---|---|---|---|---|
| United States, MS | 84 | 5 | 64 | 83 | 57 | 82 | 48 | 48.42 |
| Switzerland | 85 | 22 | 79 | 86 | 82 | 83 | 33.1 | NA |
| United States, ND | 86 | 65 | 89 | 84 | 81 | 85 | 46 | 51.32 |
| United States, AL | 87 | 9 | 84 | 87 | 76 | 88 | 47.69 | 53.37 |
| United States, MA | 88 | 87 | 86 | 89 | 88 | 86 | 48.26 | 30.89 |
| United States, NV | 89 | 15 | 87 | 88 | 83 | 89 | 45 | 22.21 |
| Brazil, AM | 90 | 89 | 90 | 90 | 89 | 90 | 60 | 36.57 |

Table S3: Ranking of disparities in the observed vaccination data based on the following metrics: final ratio of H/L, average ratio, maximum ratio over time, final difference of H-L, average difference, maximum difference over time. Gini index of wealth inequality (sources in Table S6) and Gini coefficient of vaccination are also displayed. When computing the ratio metrics, it is often the case that low SES is 0% vaccinated for several weeks while high SES has vaccinations $> 0\%$. The result is that there is asymptotic behavior of H/L in the early weeks of vaccination that does not accurately reflect the dynamics of the rollout. To avoid capturing this behavior, when calculating the ratio we excluded weeks for each country where low SES has not yet reached 0.5% vaccinated.

| Functional Response Parameters | | | | | |
|---|---|---|---|---|---|
| Name | SES | Group | Vm | Wh | k |
| Argentina | H | Sigmoidal | 1 | 26.84 | 2.75 |
| Argentina | L | Sigmoidal | 1 | 27.51 | 2.82 |
| Belgium | H | Sigmoidal | 0.82 | 19.03 | 4.08 |
| Belgium | L | Sigmoidal | 0.67 | 19.16 | 3.98 |
| Brazil | H | Sigmoidal | 1 | 21.96 | 2.82 |
| Brazil | L | Sigmoidal | 0.82 | 24.77 | 3.06 |
| Brazil, AC | H | Sigmoidal | 0.88 | 24.88 | 2.39 |
| Brazil, AC | L | Sigmoidal | 0.73 | 23.22 | 3.52 |
| Brazil, AL | H | Sigmoidal | 0.89 | 25.53 | 2.43 |
| Brazil, AL | L | Sigmoidal | 0.79 | 24.89 | 3.06 |
| Brazil, AM | H | Concave | 0.68 | 29.69 | 1.86 |
| Brazil, AM | L | Concave | 0.83 | 28.02 | 1.82 |
| Brazil, AP | H | Sigmoidal | 0.7 | 25.36 | 3.68 |
| Brazil, AP | L | Sigmoidal | 0.7 | 25.13 | 3.49 |
| Brazil, BA | H | Sigmoidal | 0.99 | 25.5 | 2.42 |
| Brazil, BA | L | Sigmoidal | 0.9 | 26.24 | 2.75 |
| Brazil, ES | H | Sigmoidal | 0.97 | 22.95 | 3.02 |
| Brazil, ES | L | Sigmoidal | 0.92 | 23.19 | 3.13 |
| Brazil, GO | H | Sigmoidal | 1 | 21.34 | 2.65 |
| Brazil, GO | L | Sigmoidal | 0.87 | 24.54 | 3.41 |
| Brazil, MA | H | Sigmoidal | 0.81 | 26.75 | 2.78 |
| Brazil, MA | L | Sigmoidal | 0.73 | 24 | 3.19 |
| Brazil, MG | H | Sigmoidal | 1 | 22.33 | 2.85 |
| Brazil, MG | L | Sigmoidal | 0.88 | 24.66 | 3.22 |
| Brazil, MS | H | Sigmoidal | 0.93 | 21.11 | 3.03 |
| Brazil, MS | L | Sigmoidal | 0.84 | 21.17 | 3.25 |
| Brazil, MT | H | Sigmoidal | 0.92 | 24.16 | 3.53 |
| Brazil, MT | L | Sigmoidal | 0.82 | 25.1 | 3.7 |
| Brazil, PA | H | Sigmoidal | 0.77 | 27.16 | 3.18 |
| Brazil, PA | L | Sigmoidal | 0.8 | 26.11 | 3.53 |
| Brazil, PB | H | Sigmoidal | 1 | 24.94 | 2.45 |
| Brazil, PB | L | Sigmoidal | 0.95 | 24.81 | 2.8 |
| Brazil, PE | H | Sigmoidal | 0.94 | 23.68 | 3.14 |
| Brazil, PE | L | Sigmoidal | 0.92 | 26.82 | 3.24 |
| Brazil, PI | H | Sigmoidal | 1 | 25.75 | 2.9 |
| Brazil, PI | L | Sigmoidal | 0.99 | 28.6 | 2.89 |
| Brazil, PR | H | Sigmoidal | 1 | 21.25 | 2.81 |
| Brazil, PR | L | Sigmoidal | 0.93 | 23.72 | 3.15 |
| Brazil, RJ | H | Sigmoidal | 0.99 | 23.09 | 2.84 |
| Brazil, RJ | L | Sigmoidal | 0.88 | 25.59 | 2.74 |
| Brazil, RN | H | Sigmoidal | 1 | 23.41 | 3.35 |

| | | | | | |
|---|---|---|---|---|---|
| Brazil, RN | L | Sigmoidal | 0.89 | 26.72 | 2.89 |
| Brazil, RO | H | Sigmoidal | 0.9 | 23.27 | 3.43 |
| Brazil, RO | L | Sigmoidal | 0.8 | 24.54 | 3.71 |
| Brazil, RS | H | Sigmoidal | 1 | 19.68 | 2.49 |
| Brazil, RS | L | Sigmoidal | 0.93 | 22.6 | 2.69 |
| Brazil, SC | H | Sigmoidal | 1 | 20.51 | 3.47 |
| Brazil, SC | L | Sigmoidal | 0.97 | 25.21 | 3.29 |
| Brazil, SE | H | Sigmoidal | 0.92 | 26.06 | 3.02 |
| Brazil, SE | L | Sigmoidal | 0.87 | 25.46 | 3.39 |
| Brazil, SP | H | Sigmoidal | 0.75 | 21.77 | 2.85 |
| Brazil, SP | L | Sigmoidal | 0.64 | 23.76 | 3.67 |
| Brazil, TO | H | Sigmoidal | 0.87 | 25.24 | 3.07 |
| Brazil, TO | L | Sigmoidal | 0.78 | 24.75 | 3.65 |
| Cape Verde | H | Concave | 0.68 | 0.95 | 0.8 |
| Cape Verde | L | Concave | 0.55 | 2.01 | 1.03 |
| Chile | H | Concave | 1 | 16.78 | 1.09 |
| Chile | L | Concave | 1 | 20.22 | 1.11 |
| Czech Republic | H | Sigmoidal | 0.7 | 20.11 | 3.58 |
| Czech Republic | L | Sigmoidal | 0.58 | 19.06 | 3.74 |
| Denmark | H | Sigmoidal | 0.82 | 19.81 | 3.8 |
| Denmark | L | Sigmoidal | 0.85 | 18.89 | 3.68 |
| Ecuador | H | Sigmoidal | 1 | 24.49 | 3.05 |
| Ecuador | L | Sigmoidal | 0.79 | 30.57 | 3.87 |
| Estonia | H | Concave | 0.68 | 18.87 | 2.49 |
| Estonia | L | Concave | 0.64 | 19.34 | 2.33 |
| Finland | H | Sigmoidal | 0.83 | 20.36 | 3.07 |
| Finland | L | Sigmoidal | 0.82 | 20.52 | 3.37 |
| France | H | Sigmoidal | 0.8 | 21.5 | 3.71 |
| France | L | Sigmoidal | 0.78 | 21.55 | 3.23 |
| Germany | H | Sigmoidal | 0.85 | 18.85 | 3.12 |
| Germany | L | Sigmoidal | 0.77 | 19 | 3.4 |
| India | H | Sigmoidal | 0.93 | 23.42 | 2.83 |
| India | L | Sigmoidal | 0.78 | 32.06 | 2.54 |
| Israel | H | Concave | 0.89 | 4.64 | 0.82 |
| Israel | L | Concave | 0.65 | 10.66 | 1.05 |
| Italy | H | Sigmoidal | 0.86 | 21.65 | 2.96 |
| Italy | L | Sigmoidal | 0.86 | 22.08 | 3.34 |
| Japan | H | Sigmoidal | 0.8 | 14.26 | 3.21 |
| Japan | L | Sigmoidal | 0.77 | 13.52 | 2.98 |
| Kazakhstan | H | Concave | 0.61 | 14.38 | 1.91 |
| Kazakhstan | L | Concave | 0.47 | 13.94 | 1.89 |
| Malaysia | H | Sigmoidal | 0.97 | 19.92 | 5.5 |
| Malaysia | L | Sigmoidal | 0.72 | 21.28 | 4.97 |

| | | | | | |
|---|---|---|---|---|---|
| Peru | H | Sigmoidal | 0.93 | 32.64 | 3.86 |
| Peru | L | Sigmoidal | 0.8 | 35.03 | 3.78 |
| Republic of Korea | H | Sigmoidal | 0.89 | 20.25 | 3.32 |
| Republic of Korea | L | Sigmoidal | 0.89 | 20.1 | 3.17 |
| Spain | H | Sigmoidal | 0.91 | 22.88 | 2.6 |
| Spain | L | Sigmoidal | 0.84 | 22.51 | 2.99 |
| Sweden | H | Sigmoidal | 0.82 | 22.45 | 3.46 |
| Sweden | L | Sigmoidal | 0.8 | 21.28 | 3.15 |
| Switzerland | H | Sigmoidal | 0.68 | 21.87 | 2.99 |
| Switzerland | L | Sigmoidal | 0.75 | 21.87 | 3.38 |
| UK | H | Concave | 0.94 | 13.16 | 1.9 |
| UK | L | Concave | 0.82 | 13.75 | 1.81 |
| United States | H | Concave | 0.82 | 14.88 | 2.13 |
| United States | L | Concave | 0.77 | 20.9 | 1.39 |
| United States, AK | H | Concave | 0.82 | 9.34 | 1.71 |
| United States, AK | L | Concave | 0.92 | 6.65 | 0.96 |
| United States, AR | H | Concave | 0.95 | 26.73 | 1.08 |
| United States, AR | L | Concave | 0.85 | 22.94 | 1.17 |
| United States, AZ | H | Concave | 0.82 | 14 | 1.66 |
| United States, AZ | L | Concave | 0.92 | 14.04 | 1.53 |
| United States, CA | H | Concave | 0.95 | 11.67 | 2.21 |
| United States, CA | L | Concave | 0.6 | 14.87 | 1.62 |
| United States, FL | H | Concave | 0.92 | 14.95 | 1.58 |
| United States, FL | L | Concave | 0.75 | 22.66 | 1.2 |
| United States, IA | H | Concave | 0.72 | 9.92 | 1.95 |
| United States, IA | L | Concave | 0.63 | 9.33 | 1.95 |
| United States, ID | H | Concave | 0.8 | 12.81 | 1.52 |
| United States, ID | L | Concave | 0.7 | 16.92 | 1.31 |
| United States, IL | H | Concave | 0.82 | 12.42 | 2.41 |
| United States, IL | L | Concave | 0.6 | 11.92 | 1.59 |
| United States, IN | H | Concave | 0.75 | 13.91 | 1.64 |
| United States, IN | L | Concave | 0.69 | 14.42 | 1.43 |
| United States, KS | H | Concave | 0.77 | 12.82 | 1.46 |
| United States, KS | L | Concave | 0.62 | 11.1 | 1.44 |
| United States, KY | H | Concave | 0.75 | 11.66 | 1.65 |
| United States, KY | L | Concave | 0.8 | 17.82 | 1 |
| United States, LA | H | Concave | 0.84 | 18.88 | 1.54 |
| United States, LA | L | Concave | 0.9 | 30.27 | 1.14 |
| United States, MA | H | Concave | 0.7 | 11.9 | 2.33 |
| United States, MA | L | Concave | 0.84 | 13 | 1.83 |
| United States, MD | H | Concave | 0.94 | 12.52 | 2.53 |
| United States, MD | L | Concave | 0.77 | 14.45 | 1.71 |
| United States, ME | H | Concave | 0.91 | 12.03 | 2.39 |

| | | | | | |
|---|---|---|---|---|---|
| United States, ME | L | Concave | 0.82 | 12.18 | 1.9 |
| United States, MN | H | Concave | 0.75 | 11.09 | 1.95 |
| United States, MN | L | Concave | 0.71 | 8.24 | 1.57 |
| United States, MO | H | Concave | 0.69 | 14.14 | 1.62 |
| United States, MO | L | Concave | 0.66 | 23.37 | 1.06 |
| United States, MS | H | Concave | 1 | 30.37 | 1 |
| United States, MS | L | Concave | 0.92 | 21.39 | 1.28 |
| United States, MT | H | Concave | 0.7 | 11.73 | 1.38 |
| United States, MT | L | Concave | 0.61 | 8.54 | 1.46 |
| United States, NC | H | Concave | 1 | 17.69 | 1.35 |
| United States, NC | L | Concave | 0.85 | 15.65 | 1.12 |
| United States, ND | H | Concave | 0.56 | 13.05 | 1.25 |
| United States, ND | L | Concave | 0.65 | 11.58 | 1.07 |
| United States, NJ | H | Concave | 0.98 | 12.25 | 2.02 |
| United States, NJ | L | Concave | 0.94 | 14.08 | 1.56 |
| United States, NV | H | Concave | 0.6 | 14.02 | 1.28 |
| United States, NV | L | Concave | 0.75 | 15.18 | 1.35 |
| United States, NY | H | Concave | 0.94 | 14.18 | 2.13 |
| United States, NY | L | Concave | 0.74 | 13 | 2.01 |
| United States, OK | H | Concave | 1 | 28.38 | 0.91 |
| United States, OK | L | Concave | 0.97 | 29.3 | 0.81 |
| United States, OR | H | Concave | 0.86 | 13.02 | 1.98 |
| United States, OR | L | Concave | 0.72 | 11.67 | 1.55 |
| United States, PA | H | Concave | 0.88 | 12.59 | 2.43 |
| United States, PA | L | Concave | 0.73 | 12.79 | 1.52 |
| United States, SC | H | Concave | 0.94 | 20.2 | 1.24 |
| United States, SC | L | Concave | 0.86 | 19.62 | 1.33 |
| United States, TN | H | Concave | 0.82 | 18.29 | 1.51 |
| United States, TN | L | Concave | 0.7 | 21.78 | 1.12 |
| United States, VT | H | Concave | 0.8 | 12 | 2.72 |
| United States, VT | L | Concave | 0.69 | 12.8 | 2.2 |
| United States, WA | H | Concave | 0.85 | 12.91 | 2.13 |
| United States, WA | L | Concave | 0.79 | 15.07 | 1.28 |
| United States, WI | H | Concave | 0.78 | 11.25 | 1.78 |
| United States, WI | L | Concave | 0.76 | 10.62 | 1.58 |
| United States, WY | H | Concave | 0.73 | 13.48 | 1.39 |
| United States, WY | L | Concave | 0.58 | 10.93 | 1.25 |
| Uruguay | H | Concave | 0.85 | 7.97 | 1.64 |
| Uruguay | L | Concave | 0.91 | 8.3 | 1.51 |
| Mean Parameters | H | Sigmoidal | 0.9 | 22.9 | 3.14 |
| Mean Parameters | H | Concave | 0.82 | 14.46 | 1.71 |
| Mean Parameters | L | Sigmoidal | 0.82 | 23.92 | 3.32 |
| Mean Parameters | L | Concave | 0.75 | 15.23 | 1.45 |

Table S4: Functional response parameters for each place in the fit data. Means for each group and SES level are included at the bottom. Note: Brazil and the US are included as both countries and states, but only the individual states are used to compute group averages.

| Model Parameters | | | | |
|---|---|---|---|---|
| Symbol | Description | Value | Units | References |
| $\mu$ | probability of transmission given contact | 0.42 | proportion | [77] |
| $c_{LL}$ | number of contacts | $3 \times 0.7$ | | assumption |
| $c_{LH}$ | number of contacts | $3 \times 0.7$ | | assumption |
| $c_{HL}$ | number of contacts | $1 \times 0.4$ | | assumption |
| $c_{HH}$ | number of contacts | $3 \times 0.4$ | | assumption |
| $\gamma_A$ | recovery rate (asymptomatic) | 1/5 | 1/days | [15, 78–80] |
| $\gamma_S$ | recovery rate (symptomatic) | 1/10 | 1/days | [78–80] |
| $\epsilon$ | latent period | 1/3 | 1/days | [15, 81] |
| $a_{L1}$ | proportion mild/asymptomatic | 0.5 | proportion | assumption |
| $a_{H1}$ | proportion mild/asymptomatic | 0.5 | proportion | assumption |
| $a_{L2}$ | proportion mild/asymptomatic | 0.7 | proportion | [71] (upper bound) |
| $a_{H2}$ | proportion mild/asymptomatic | 0.7 | proportion | [71] (upper bound) |
| $\omega$ | rate of waning immunity | 1/120 | 1/days | assumed |
| $b$ | scale waning immunity for vaccinated | 0.5 | scalar value | computed from $\omega$ and need for booster after 8 months |
| $ss$ | scale susceptibility for class 2 | 0.5 | scalar value | assumption |
| $si$ | scale infectiousness for class 2 | 0.5 | scalar value | assumption |
| $\alpha_{H1}$ | infection fatality rate | 0.01 | proportion | [2] |
| $\alpha_{H2}$ | infection fatality rate | 0.00178 | proportion | [2, 72, 73] |
| $\alpha_{L1}$ | infection fatality rate | 0.028 | proportion | [2] |
| $\alpha_{L2}$ | infection fatality rate | 0.00485 | proportion | [2, 72, 73] |

Table S5: Parameters used in model simulations.

| Gini/Lorenz Sources | | |
|---|---|---|
| Location | Type | Source |
| Global | Gini coefficient (figure S8) | World Population Review |
| Global | Gini coefficient (figure S8) | World Population Review |
| Cape Verde | Gini coefficient (figure S8) | World Bank |
| Brazilian states | Gini coefficient (figure S8) | do Socorro Candeira Costa et al., 2017 |
| United States | Population (2019) (for Lorenz Curve) | Github (Census) |

Table S6: Additional data sources for Lorenz and Gini coefficient analysis.

### S3 Supplementary Figures

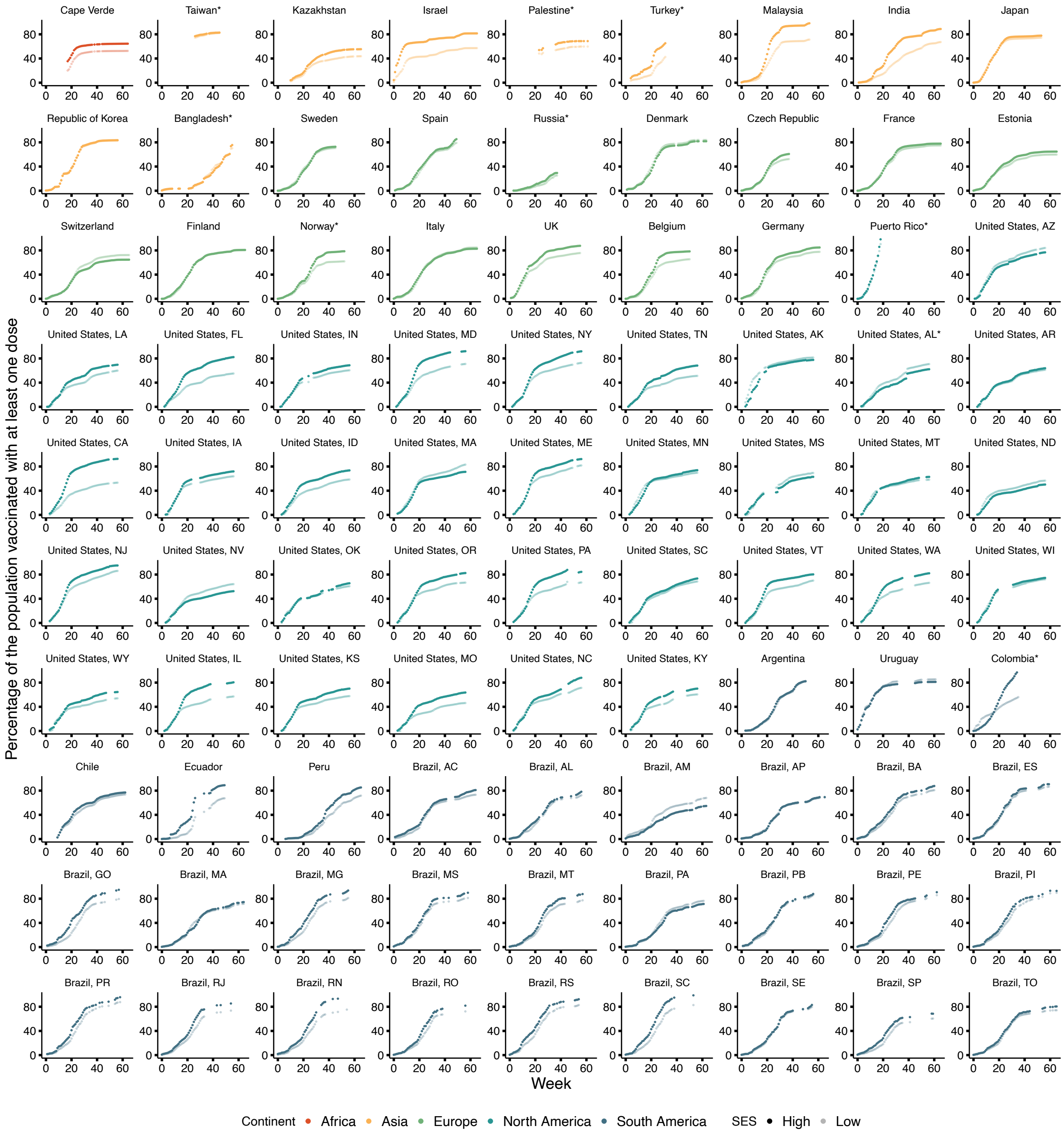


Figure S1: Observed data for all countries and territories analyzed. Data for Colombia and Puerto Rico do not differentiate between the number of doses, so these specific plots show the total number of doses administered.

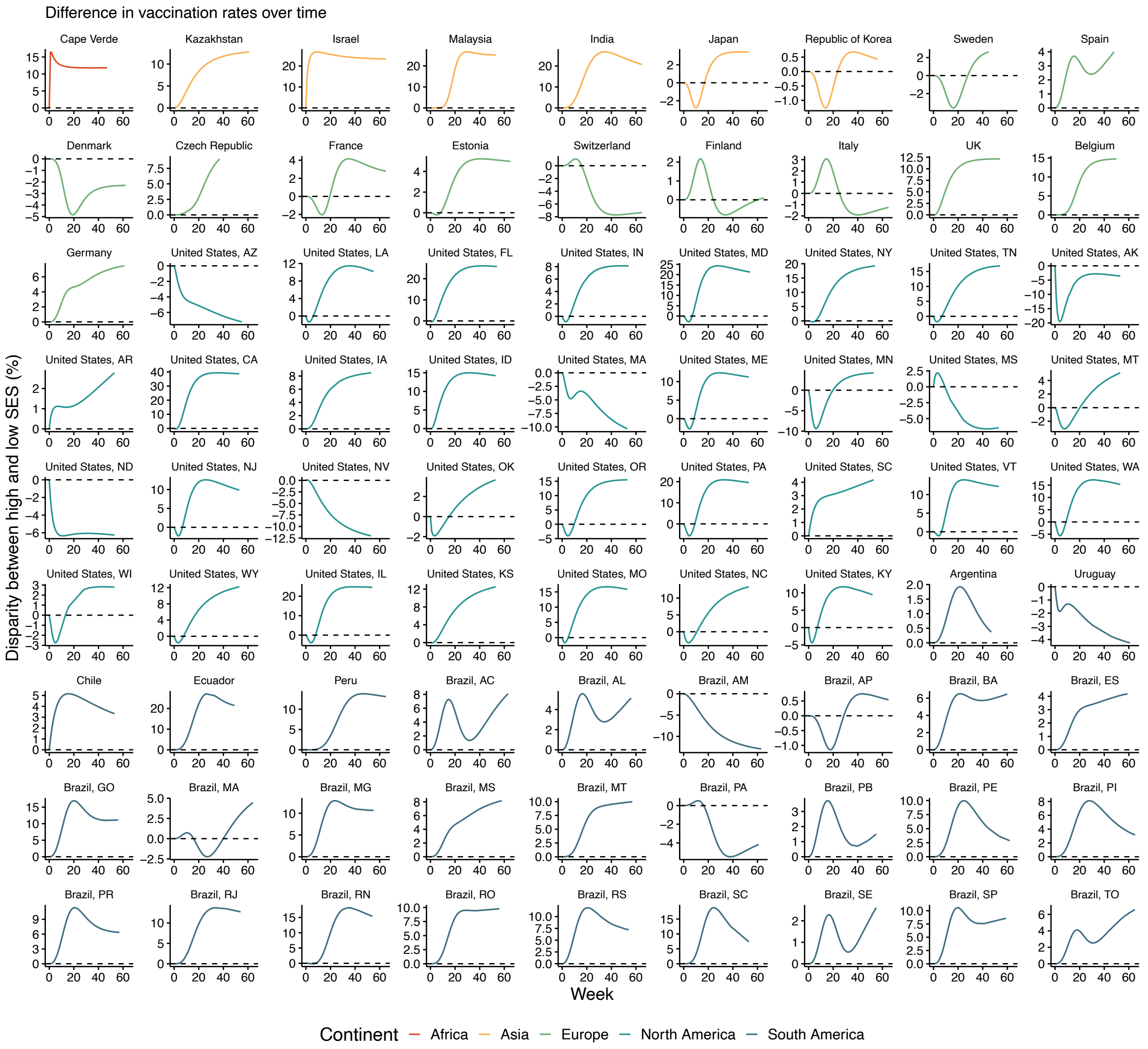


Figure S2: Vaccine disparity over time based on the fit data. We take the difference (H-L) of the vaccine coverage. The curves for each country are colored by continent.

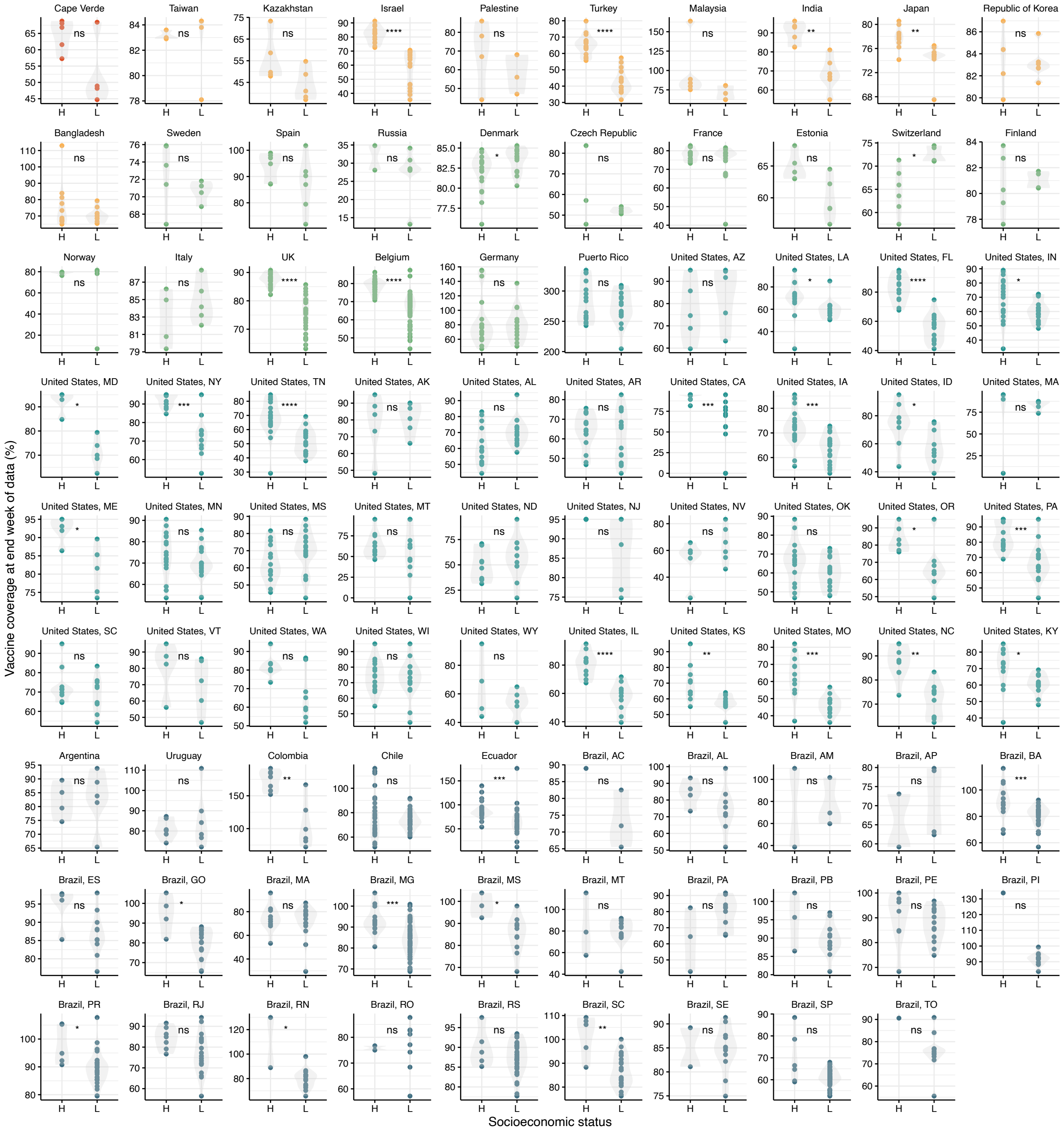


Figure S3: Comparison of location-specific vaccination % in the final week of the data, grouped by socioeconomic status and country. There is a disparity in favor of high SES for over 80% of the places analyzed. P-values are shown.

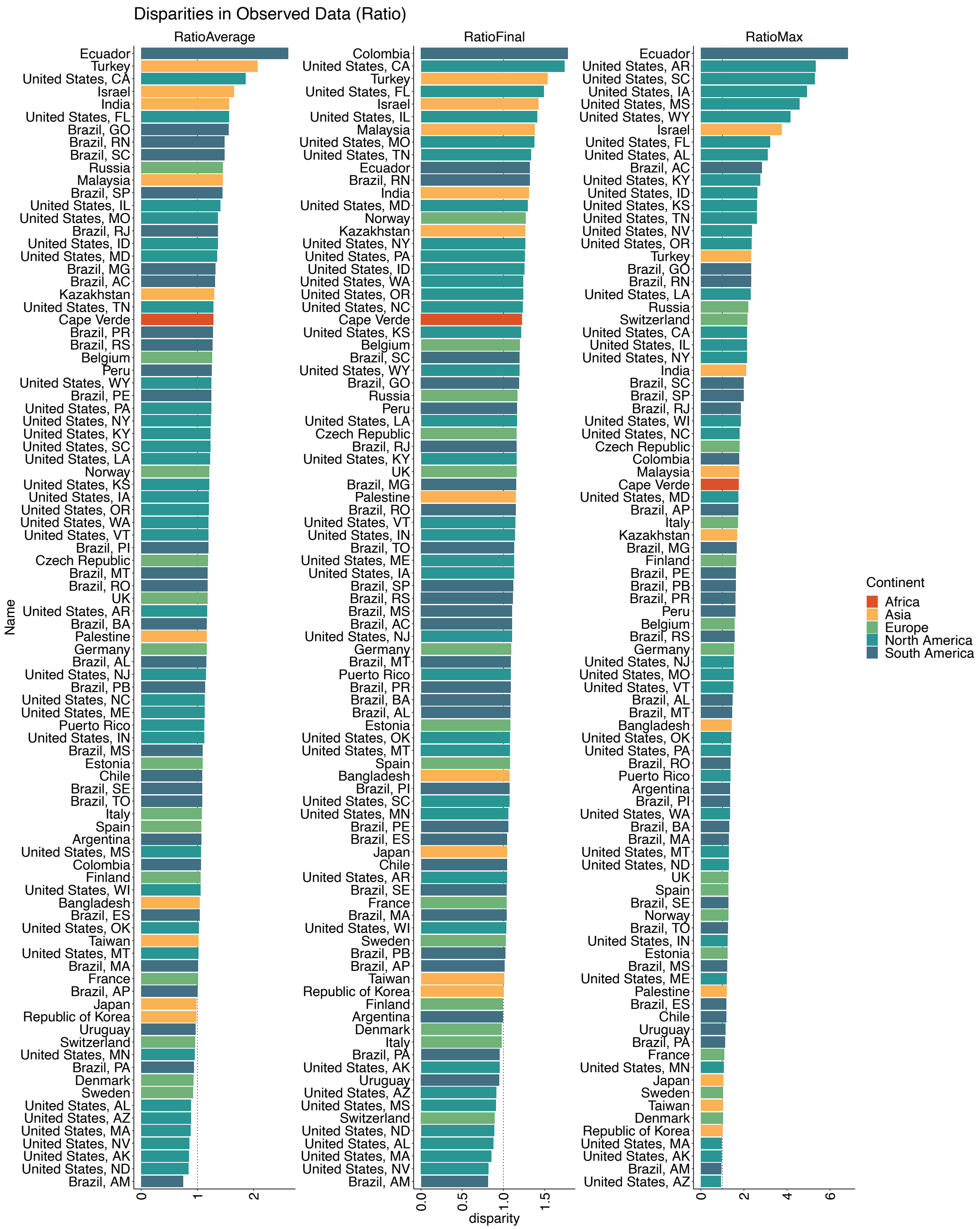


Figure S4: Bar plot of disparities in vaccination between low vs. high SES. Calculated as a ratio (H/L). When computing the ratio metrics, it is often the case that low SES is 0% vaccinated for several weeks while high SES has vaccinations > 0%. The result is that there is vertically asymptotic behavior of H/L in the early weeks of vaccination that does not accurately reflect the dynamics of the rollout. To avoid capturing this behavior, when calculating the ratio we excluded weeks for each country where low SES has not yet reached 0.5% vaccinated.

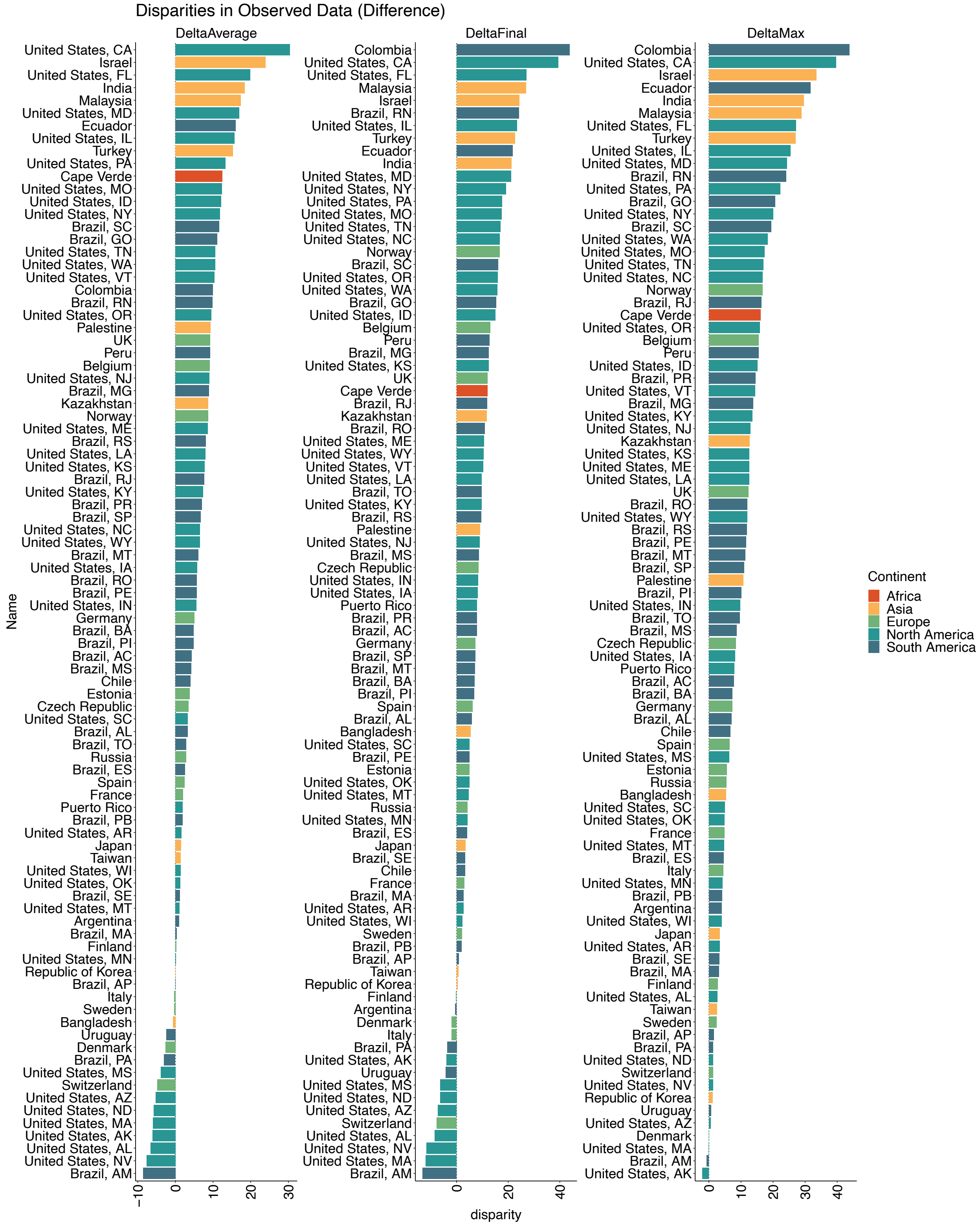


Figure S5: Bar plot of disparities in vaccination between low vs. high SES. Calculated as a difference (H-L).

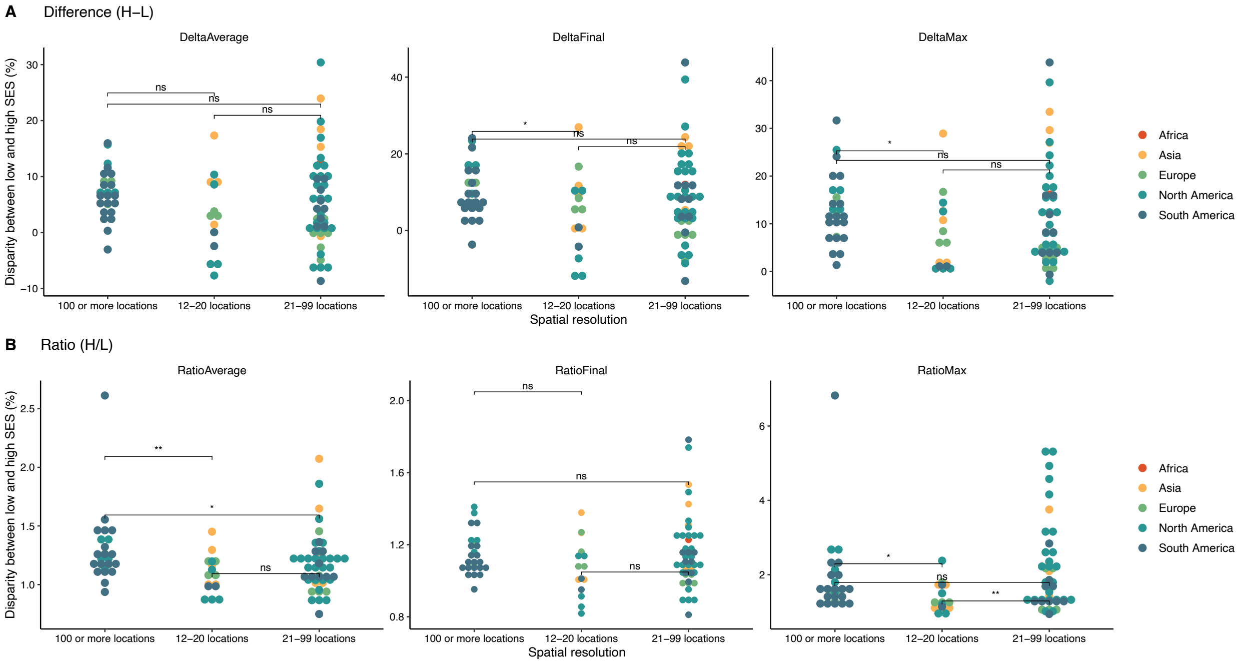


Figure S6: Analysis of disparity associations with spatial resolution, computed as a difference (A) or ratio (B).

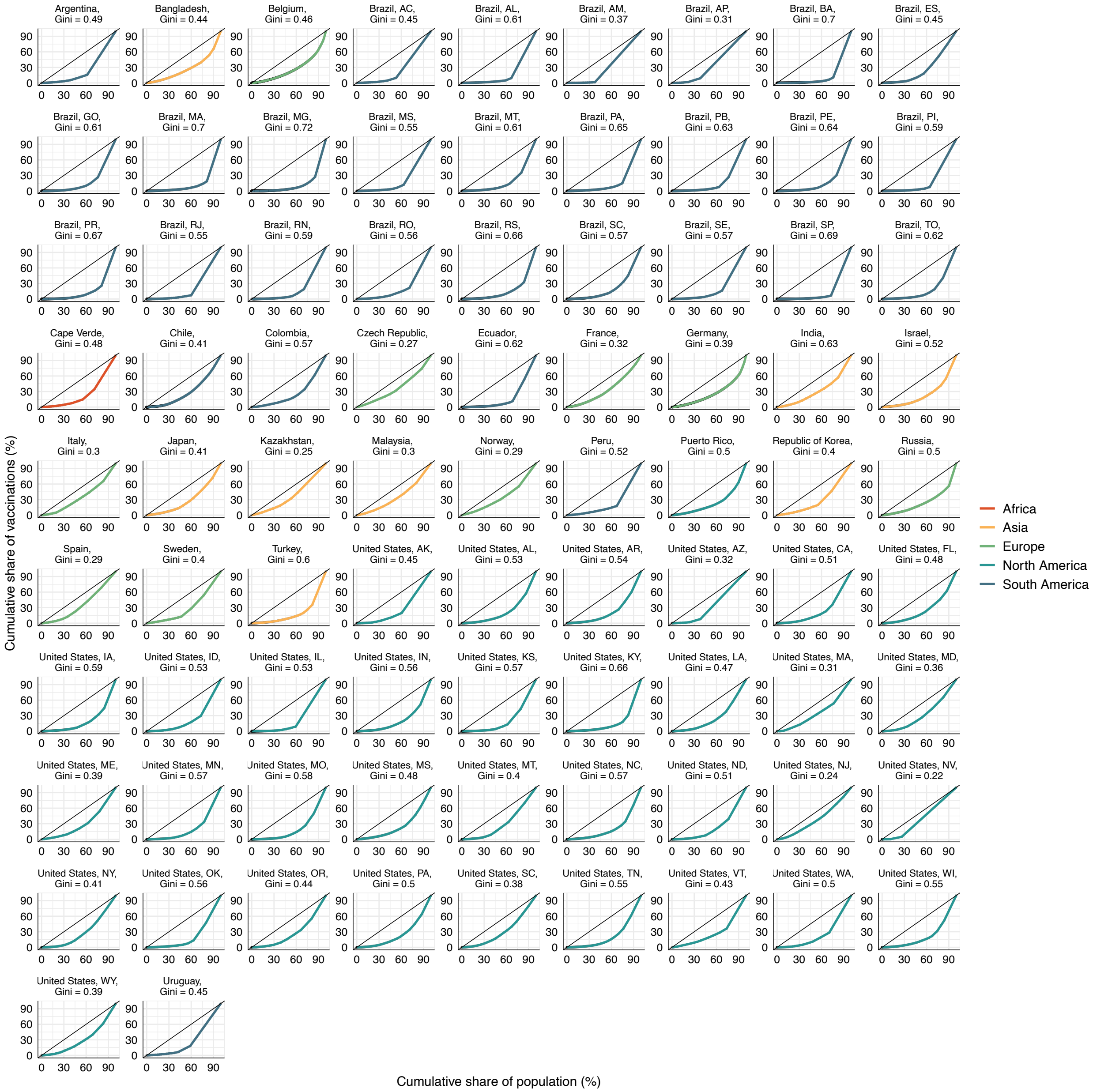


Figure S7: Lorenz curves for 83 countries, territories, and states. We filtered for the maximum week reported in the data and analyzed cumulative vaccination against share of the population. All data is for the first-dose except for Puerto Rico, which reports cumulative vaccinations (all types). Each point on the curve can be read as "X% of the population holds Y% of the vaccination doses". Inequity is seen in every country and state, to varying degree.

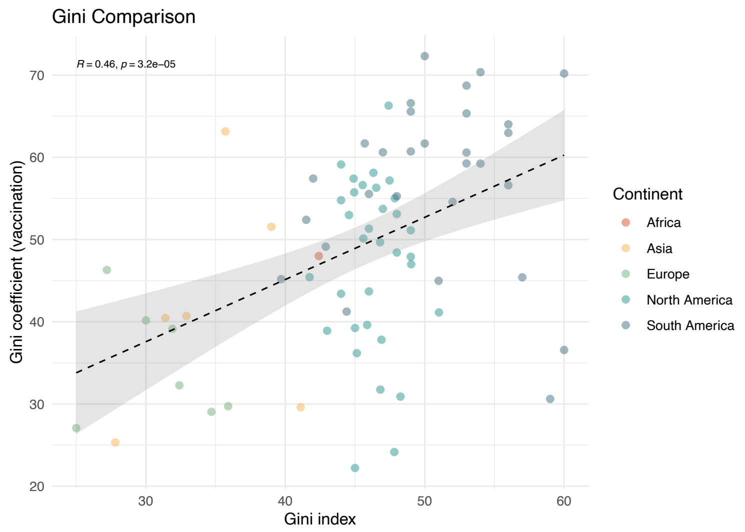


Figure S8: Comparison of Gini index of income distribution and of vaccination for $n = 52$ places. The degree of income inequality in a country is significantly associated with the degree of vaccine disparity.

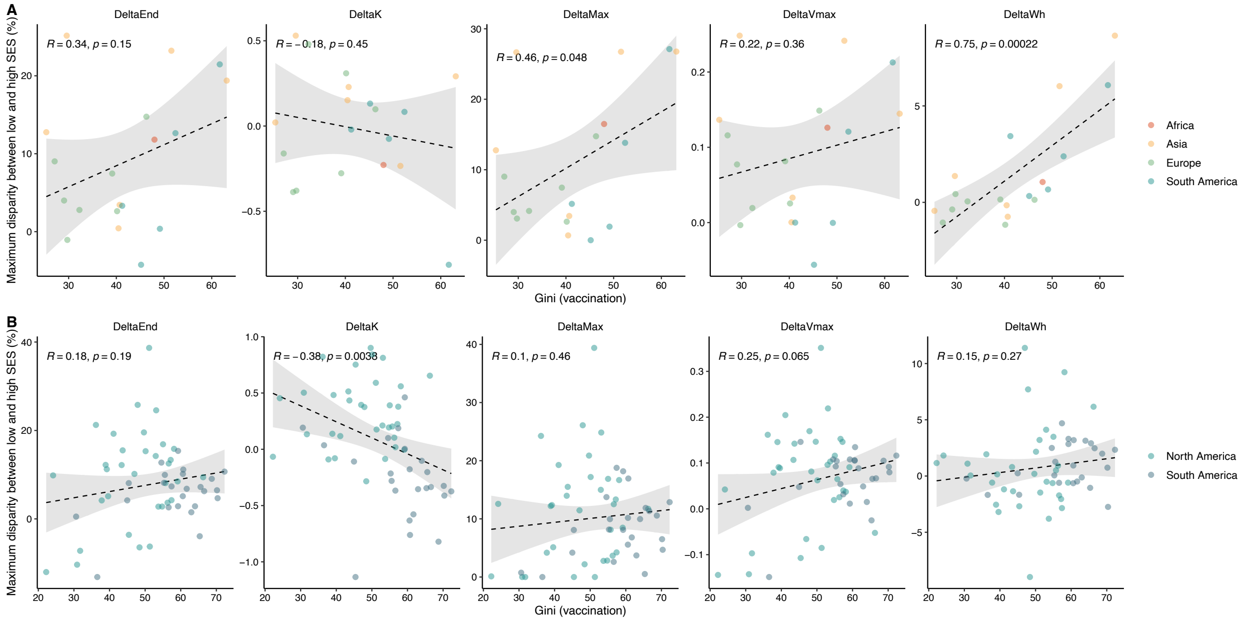


Figure S9: Based on the Gini coefficient of the Lorenz curves of vaccine distribution, we analyze 83 countries and US and Brazilian states - countries are shown in panel (A) and states in panel (B). DeltaEnd is the % disparity between high and low SES at the end of the predicted data, whereas DeltaMax is the maximum the disparity ever reaches in the predicted data. DeltaK, DeltaVmax, and DeltaWh are the disparities in the corresponding functional response parameters between high and low SES. At the state level, DeltaK is significantly negatively associated with the Gini coefficient of vaccination. At the country level, DeltaMax and DeltaWh are significantly positively associated.

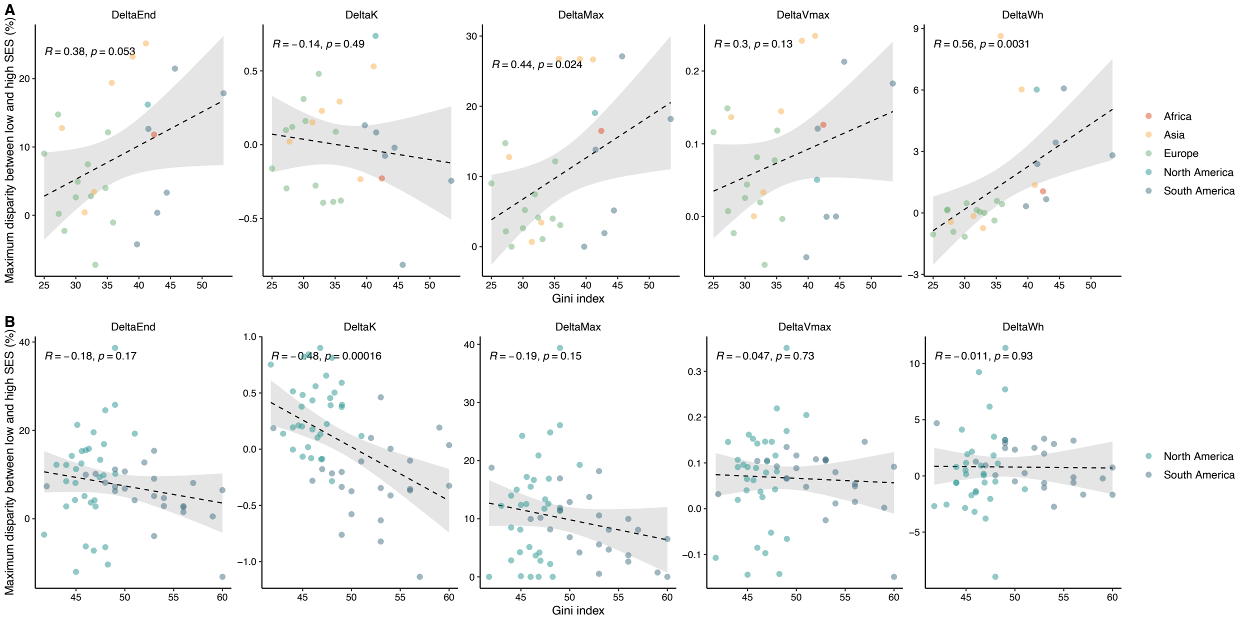


Figure S10: Gini index analysis for 59 places - countries are shown in panel (A) and US and Brazilian states in panel (B). DeltaEnd is the % disparity between high and low SES at the end of the predicted data, whereas DeltaMax is the maximum the disparity ever reaches in the predicted data. DeltaK, DeltaVmax, and DeltaWh are the disparities in the corresponding functional response parameters between high and low SES. At the state level, DeltaMax is significantly negatively associated with the Gini index. At the country level, DeltaMax and DeltaWh are significantly positively associated with the Gini index.

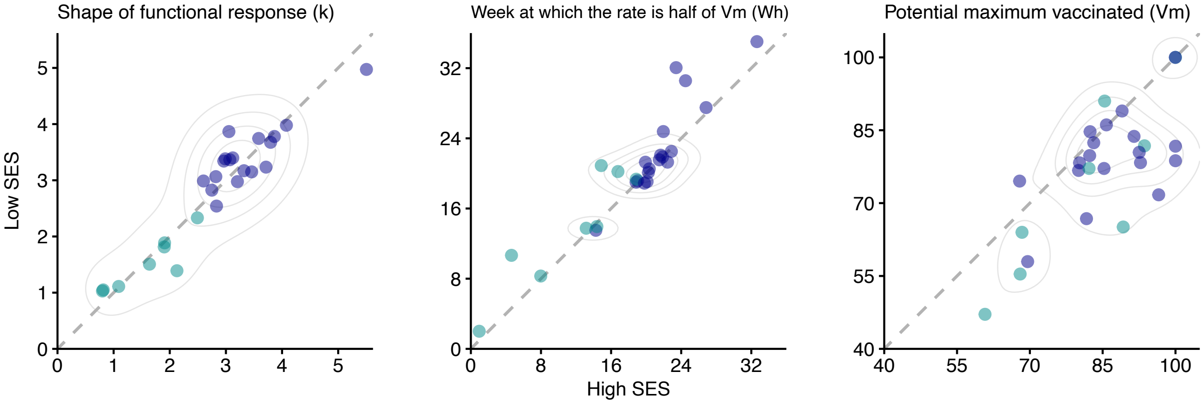


Figure S11: Alternate version of fig. 3. where the US and Brazil are displayed as whole countries rather than including individual states. While the clustering is weaker, there are still two distinct groups.

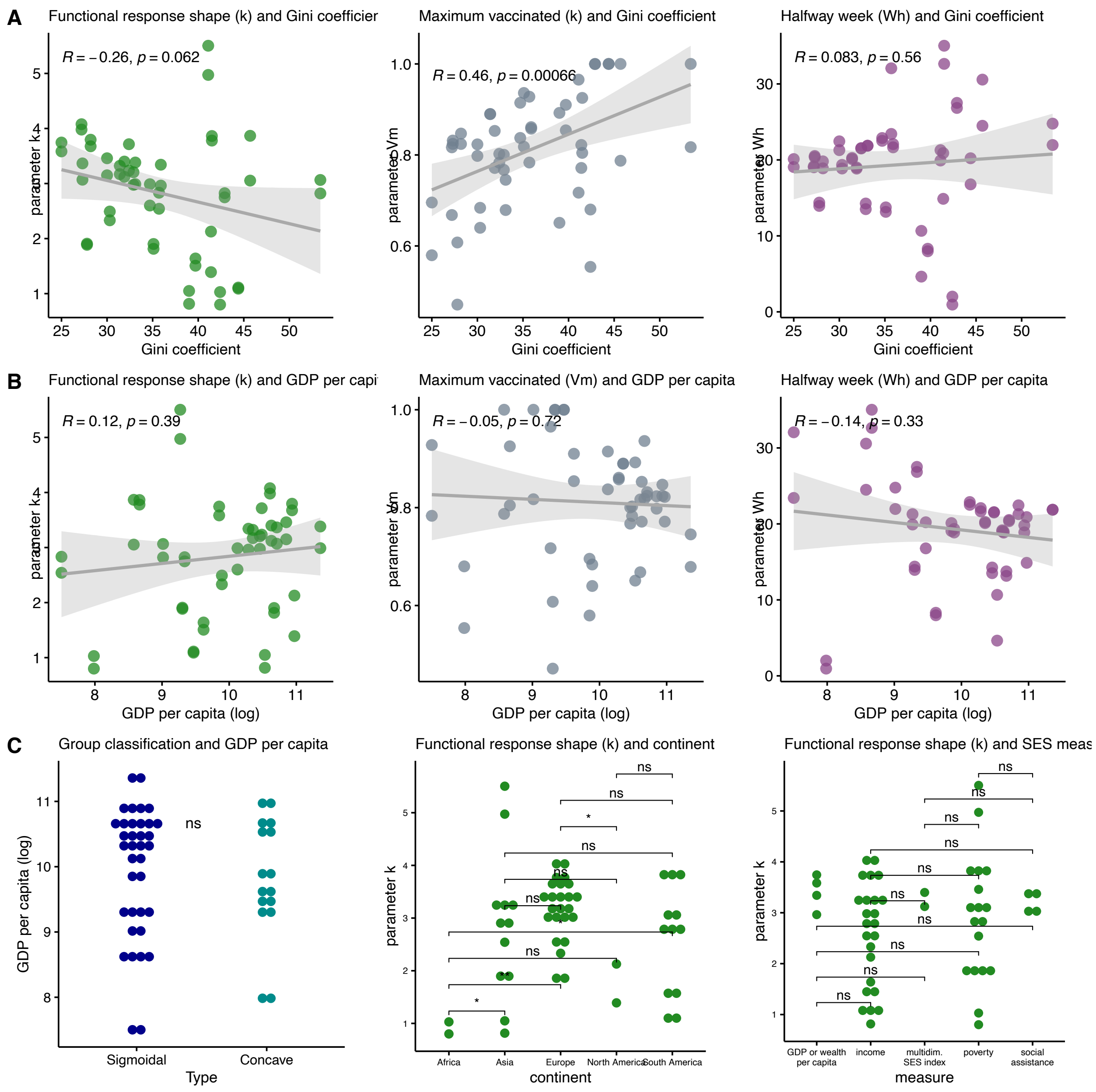


Figure S12: Analysis of parameter associations. (A) Within the subnational locations, only Vm is significantly associated with the Gini coefficient of income distribution. (B) Within the subnational locations, none of k, Vm, or Wh is significantly associated with GDP per capita. (B) Trend classification is not associated with GDP, and functional response (k) is largely unassociated with continent or measure selected.

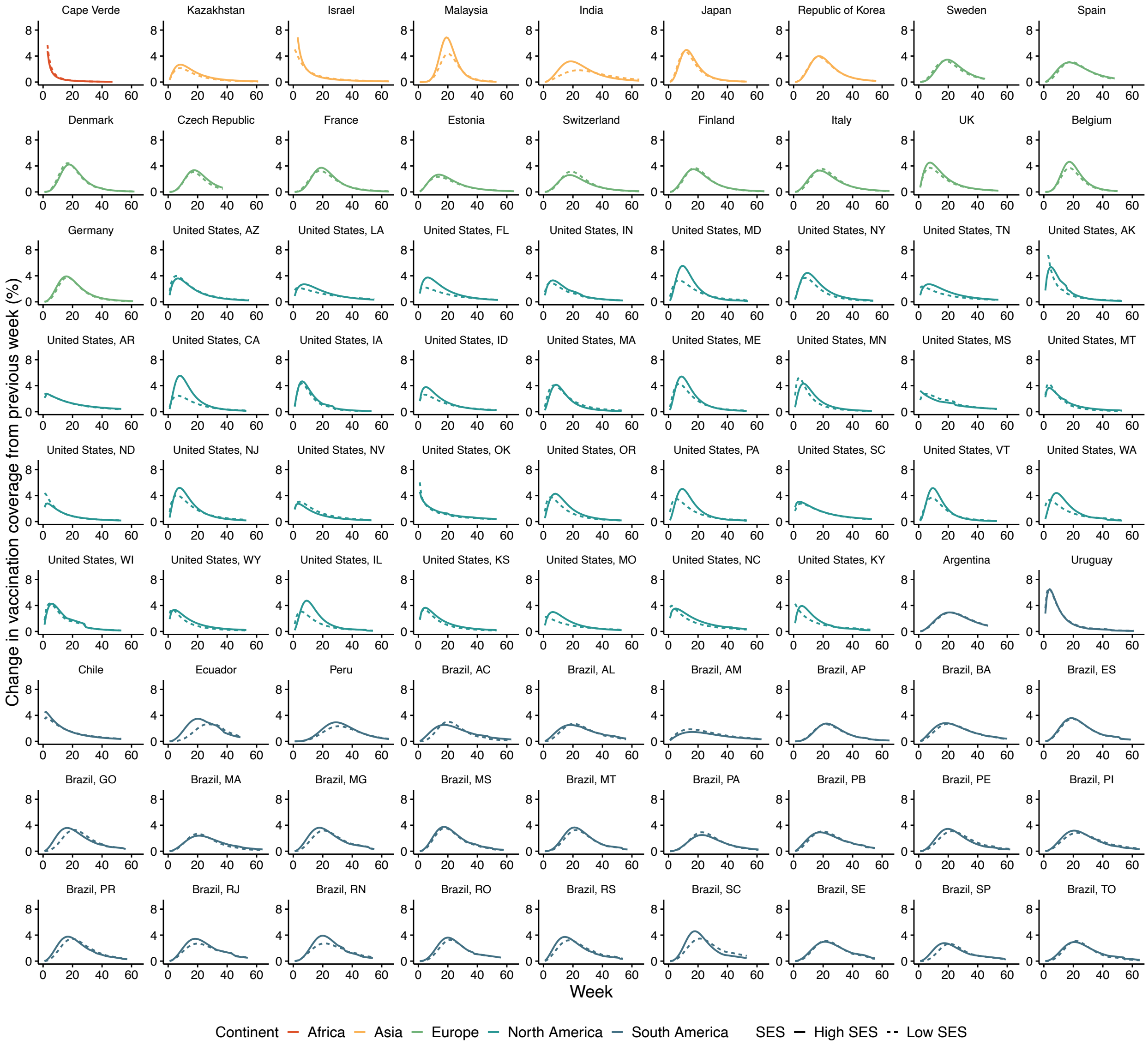


Figure S13: Estimate of the 1st derivative of vaccination trends over time, based on the fit data, for high (solid) and low (dashed) SES.

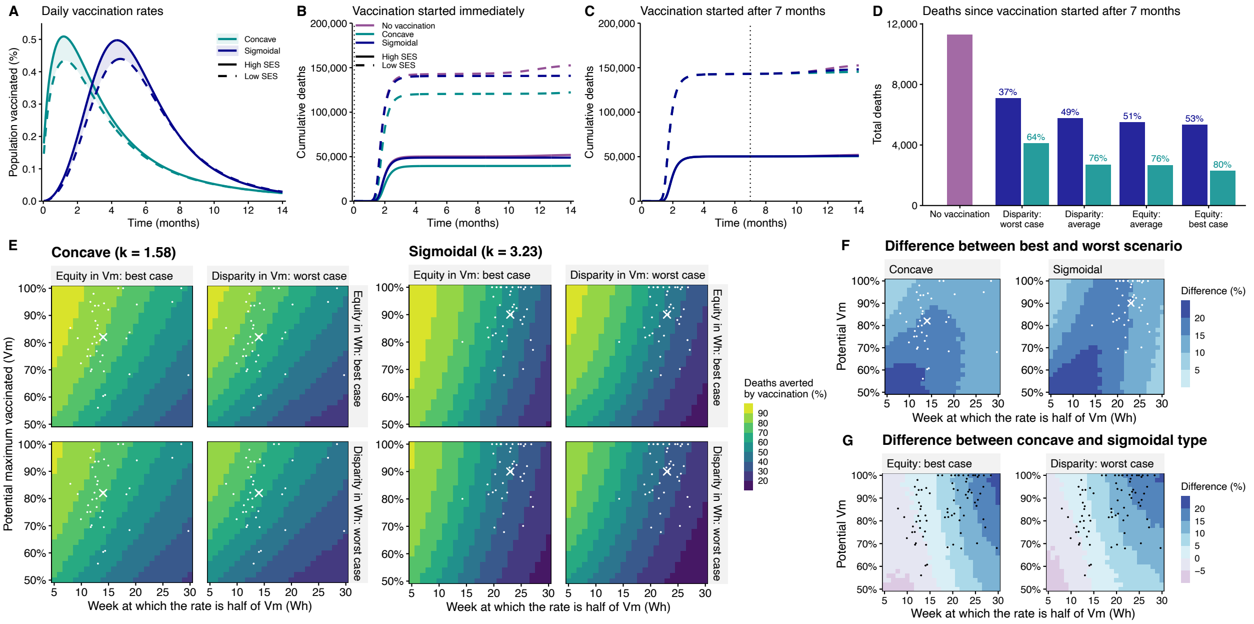


Figure S14: Analysis of deaths under different vaccination rollout scenarios. (A) Average daily vaccination rates for each type and SES group (per-country rates shown in fig. S13). In the sigmoidal type, the parameters for high SES are $Vm = 89\%$ and $Wh = 23$ weeks, while for low SES are $Vm = 82.5\%$ and $Wh = 24$ weeks. For both SES groups, $k = 3.23$, the average across SES. In the concave type, the parameters for high SES are $Vm = 82.2\%$ and $Wh = 14$ weeks while for low SES they are $Vm = 75.5\%$ and $Wh = 15$ weeks. For both SES groups, $k = 1.58$. (B) Cumulative deaths per 10,000 with vaccination starting immediately. (C) Cumulative deaths per 10,000 with vaccination starting at $t = 7$ months. (D) Cumulative deaths per 10,000 after 14 months under 4 scenarios of sociocioeconomic disparity: disparity: average, disparity: worst case (95th percentile), equity: average (averaging high SES and low SES parameters), and equity: best case (assuming vaccination of the low SES group follow the same parameters as the high SES group. (E) Deaths averted by vaccination. Keeping $k = 1.58$ for concave and $k = 3.23$ for sigmoidal, we fixed the low SES parameters at a constant disparity from high SES based on the 95th percentiles of the observed data ($\Delta Vm = 21\%$, $\Delta Wh = 6$ weeks), or considered low SES parameters equal to high SES (equity). The effect of varying parameter $k$ can be found in figure S17. (F) The difference in deaths averted under the best-case equity scenario and worst-case disparity, for both concave and sigmoidal types. (G) The difference in deaths averted under concave versus sigmoidal types, for both best-case equity and worst-case disparity. Average high SES parameters are illustrated with a cross, while country-specific parameters for high SES are shown with dots. Model parameters are listed in table S5.

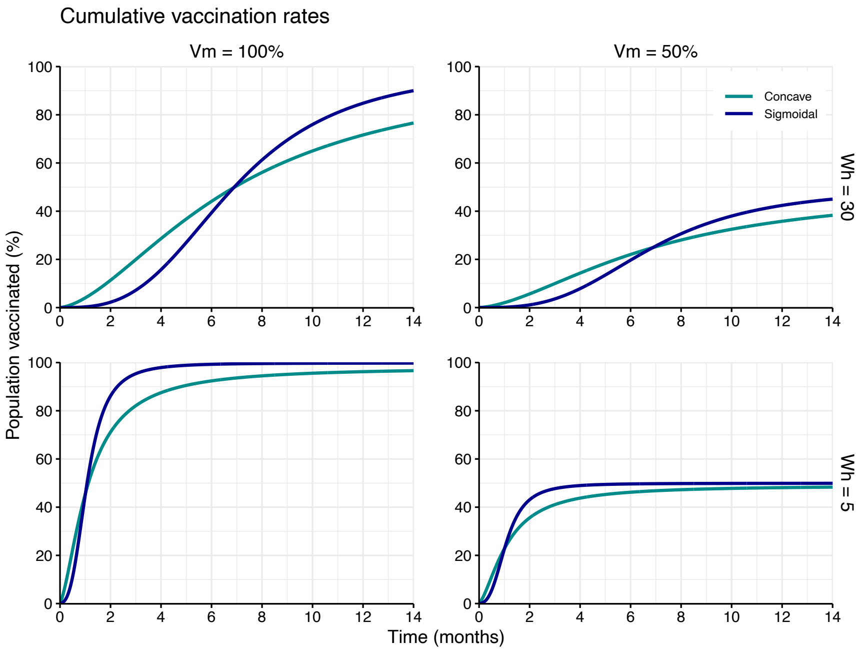


Figure S15: Cumulative vaccination trends under extreme conditions (Vm = 50 or 100%, Wh = 5 or 30 weeks).

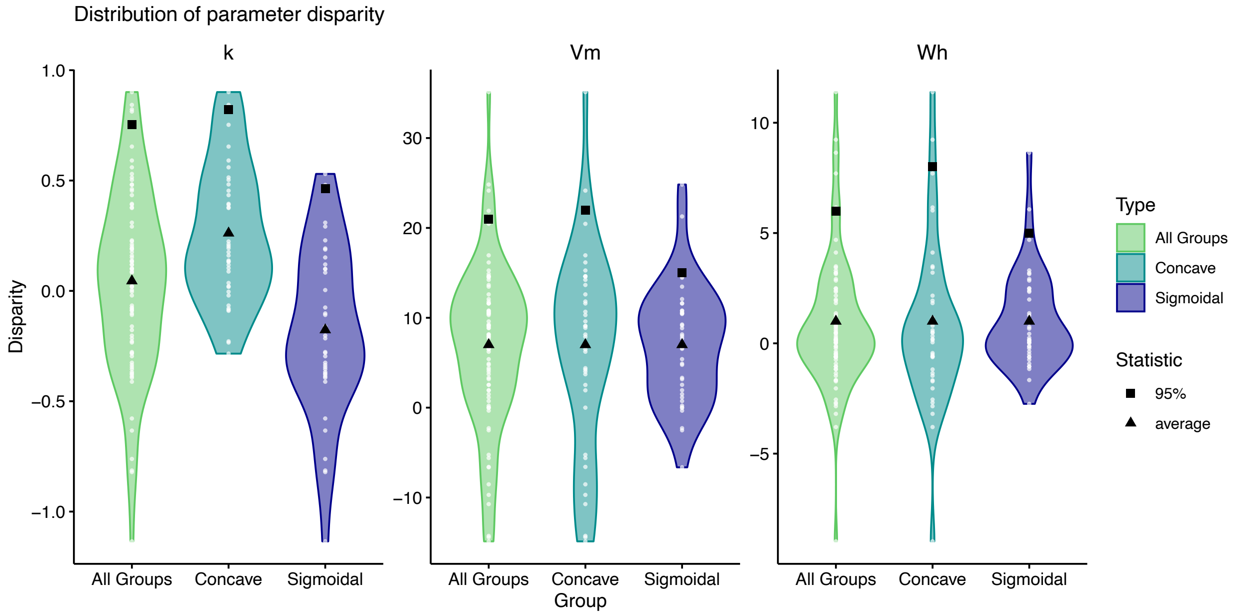


Figure S16: Distribution of functional response parameter disparities between high and low SES, either by type (concave vs. sigmoidal) or aggregated (all groups), with the average (triangle) and 95th percentile (square).

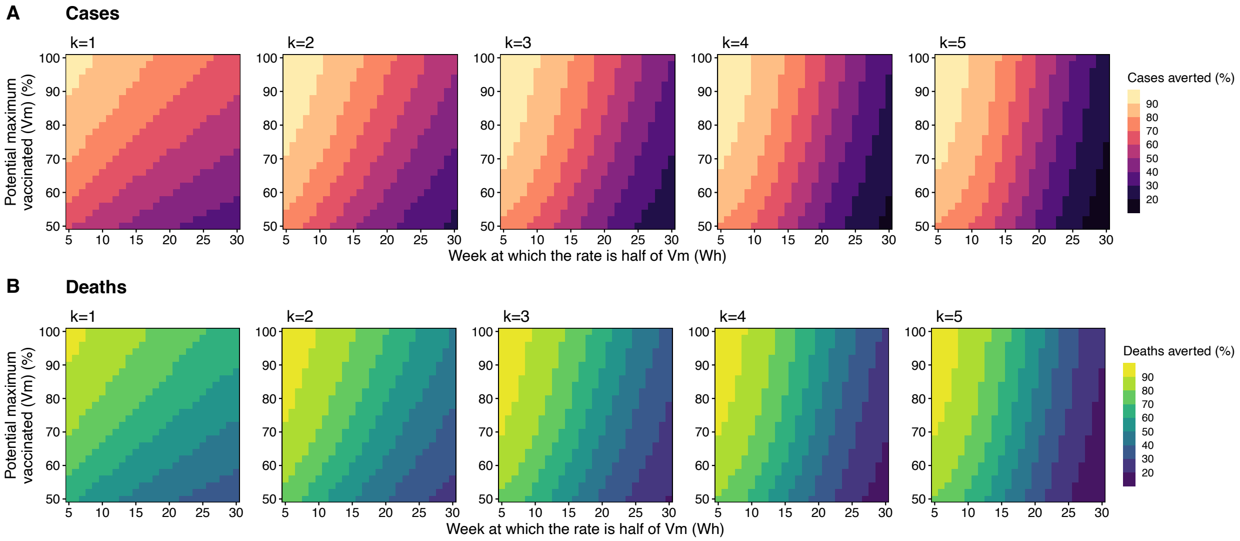


Figure S17: Cases and deaths averted for the average disparity scenario (delta $W_h$ = 1 week, delta $V_m$ = 7%) for 5 different values of $k$. Full set of the parameters can be found in table S5.

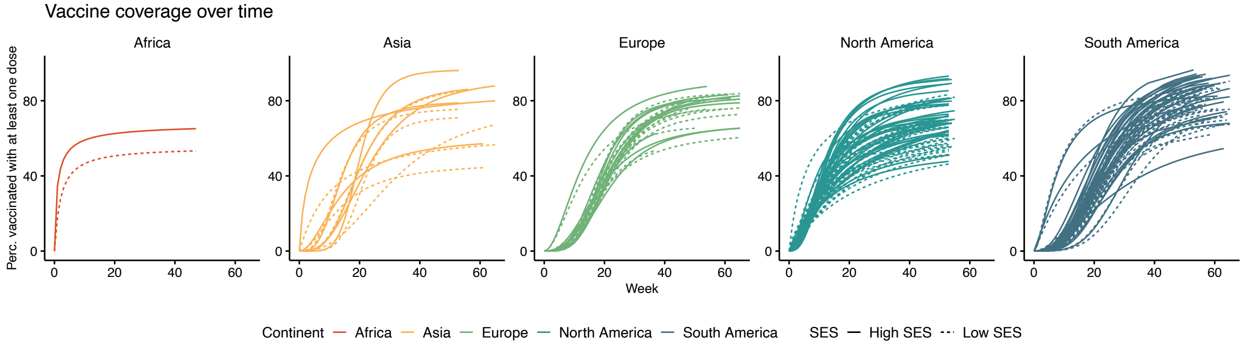


Figure S18: Cumulative percentage of the population vaccinated with at least one dose over time, colored and grouped by continent. High and low SES are shown in solid and dashed lines, respectively.
